# Health Disparities in Complicated Lower Respiratory Tract Infections: a population-based cohort study in The Hague, The Netherlands

**DOI:** 10.1101/2025.06.23.25330102

**Authors:** Ernst D. van Dokkum, Martijn Sijbom, Saskia Le Cessie, Dennis O. Mook-Kanamori, Marc A. Bruijnzeels, Adriënne S. van der Schoor, Karlijn te Paske, Kirsten Langeveld, Leo G. Visser, Mattijs E. Numans, Cees van Nieuwkoop, Hanneke Borgdorff

## Abstract

**Introduction:** International studies show that ethnic minority groups and individuals with lower socioeconomic status (SES) face higher severity of lower respiratory tract infections (LRTI). It is unclear whether these associations exist in the Netherlands and to what extent they are mediated through comorbidities and household composition. Our study investigates the relationship between sociodemographic factors and complicated LRTI before and during the COVID-19 pandemic in The Hague, the Netherlands.

**Methods:** A population-based cohort study was conducted using sociodemographic and insurance claims data from Statistics Netherlands, covering The Hague’s adult population in 2014-2019 (pre-COVID-19) and 2020 (COVID-19). The associations of structural determinants (age, sex, SES, migration background) and intermediary determinants (comorbidities, household composition) on complicated LRTI incidence and mortality were assessed using Poisson regression models. Complicated LRTI episodes were defined as hospital admission or death within 14 days after an emergency department visit.

**Results:** In 2014-2019, individuals in the lowest SES quintile displayed the highest incidence of complicated LRTI (aIRR 2.26 [95%CI:1.98–2.57]), and LRTI-associated mortality (aIRR 3.49 [95%CI:2.28– 5.34]), compared to the highest SES quintile, after adjusting for structural and intermediary determinants. Similar associations were observed in 2020. Individuals without a migration background were most affected in 2014-2019, while individuals with a migration background were more impacted in 2020.

**Conclusions:** Lower SES was consistently associated with a higher incidence of complicated LRTI. Populations with a migration background were particularly vulnerable during the pandemic. These findings underscore the need to consider SES and migration background in targeted treatment, prevention, and population health strategies.

**Summary Box:** *What is already known on this topic:* Studies from the United States and United Kingdom have shown that area deprivation, socioeconomic status (SES) and/or migration background are associated with a complicated course of lower respiratory tract infections (LRTI). However, the generalisability to of these findings to other countries is unclear due to differences in population composition and health systems. Furthermore, several factors mediate in the association between SES, migration background and complicated LRTI. However, their relative importance remains insufficiently understood and studies often do not take these mediating factors into account. Therefore, SES and migration background are currently rarely incorporated into European clinical or public health guidelines for LRTI prevention or treatment.

*What this study adds:* This study finds consistent associations between socioeconomic status (SES) and complicated LRTI both before and during the COVID-19 pandemic in an urban setting in The Netherlands, while migration background was strongly associated with complications during the first year of the pandemic. These associations were only partly mediated by comorbidities and household composition. The study also identified geographical hotspots concerning complicated LRTI incidence in The Hague, which overlapped with neighbourhoods of greater socioeconomic deprivation.

*How this study might affect research, practice or policy:* Current primary care and public health guidelines for LRTI prevention and treatment focus on clinical factors such as age and comorbidity, while overlooking the residual effect of SES and migration background. The data presented in this study, highlights significant health disparities in complicated LRTI, with area deprivation, lower SES and migration background strongly associated with complicated lower respiratory tract infections. This underscores the need for further research into other mediating factors and targeted interventions, such as incorporating SES as a risk factor in clinical decision making and focusing population- and public health strategies on deprived areas.

## Introduction

Lower respiratory tract infections (LRTI) are the fourth leading cause of disability-adjusted life-years globally and the most common infectious cause of mortality in Europe [1]. In the Netherlands, prior to the COVID-19 pandemic, approximately 530,000 patients developed a LRTI annually, of whom approximately 50,000 required hospitalisation each year [2, 3].

Risk factors for a complicated course of LRTI include advanced age, smoking history, lack of vaccination, and comorbidities such as diabetes mellitus, cancer, chronic cardiovascular, respiratory, liver, or renal disease, and immunosuppression [4–6]. These risk factors were further substantiated during the COVID-19 pandemic when hospitalisation and mortality risk were linked to age, vaccination status, obesity, and comorbidities [6, 7]. Therefore, current LTRI prevention strategies (e.g., vaccination campaigns) in Europe are focused mainly on older adults and individuals with comorbidities [8].

Studies from the United States and the United Kingdom suggest that ethnic minority groups [9, 10], and individuals in areas with higher deprivation and lower socioeconomic status (SES) also face higher LRTI hospitalisation and mortality rates [11–14]. However, the generalisability of these findings to the Dutch context is unclear due to differences in healthcare systems, population composition and environmental factors [15]. Furthermore, it remains unclear whether sociodemographic determinants are similar for pre-COVID epidemic LRTI and the COVID-19 pandemic. While a few Dutch studies have linked migration background and lower SES regions to complicated COVID-19 [16–19], data on other LRTI are lacking.

SES and migration background influence LRTI outcomes through mediators such as healthcare access, health literacy, psychological factors, lifestyle, living conditions, environmental exposures, and comorbidities [11, 20]. However, their relative contributions remain insufficiently understood. The ‘WHO Framework of Social Determinants of Health’ provides a valuable framework to study these contributions, distinguishing between structural determinants (e.g., SES) and intermediary determinants (e.g., behaviours) affecting health outcomes [21].

A better understanding of the extent to which health disparities are present in the population in complicated LRTI is essential for designing targeted population health interventions and public health policy. As more than half of the population of The Hague has a migration background and The Hague has the highest income-based segregation in The Netherlands, it provides an ideal setting to explore neighbourhood-level disparities in complicated LRTI incidence [22]. This study aimed to identify health disparities in complicated LRTI in The Hague, The Netherlands, based on socioeconomic area deprivation, SES and migration background, while also evaluating the extent of health disparities and the role of clinical mediators.

## Methods

### Study Design and Subjects

A population-based cohort study was conducted based on non-public pseudonymised data from Statistics Netherlands (SN) from January 1^st^, 2014, to December 31^st^, 2020, covering the adult population (≥18 years) of the city of The Hague (n ≈ 420,000 annually). Data are collected by SN from various administrative sources linked to the Dutch population register, including sociodemographic, hospital claims, and pharmacy claims data. The incoming data are pseudonymised by SN and made available within a secure environment to the Extramural LUMC Academic Network (ELAN) data infrastructure [23, 24]. Through this mechanism, data on age, sex, SES, migration background, household composition, medication, hospital admissions, diagnoses, mortality, and a 4-digit postal code were obtained.

### Outcome Measures

Our primary outcome, a complicated LRTI case, was defined as a patient with a pneumonia or influenza-like illness (ILI) diagnosis, who visited the emergency department (ED), received chest imaging (X-ray or Computed Tomography) and was subsequently admitted to the hospital or died within 14 days. Furthermore, to focus solely on community-acquired cases, patients discharged from a hospital within 30 days before the ED visit were excluded to avoid including hospital-acquired cases. Those who died with LRTI as a registered cause of mortality, irrespective of an ED visit, were also considered complicated LRTI cases.

Our secondary outcome, LRTI-associated mortality, was defined as a LRTI patient who died within 30 days from the ED visit or as an individual who died with LRTI as a registered mortality cause, irrespective of an ED visit.

### Determinants

A four-digit postal code was used to determine in which city neighbourhood an individual resided at the time of observation [26].

The following determinants were included as covariates: 1) age; 2) sex assigned at birth; 3) household SES defined as financial prosperity using a composite measure of the household’s standardised annual disposable income and wealth [25]; 4) migration background determined by an individual’s country of birth when born abroad or the parent’s country of birth when the individual was born in The Netherlands [25]; 5) household composition based on the number of adults and children living in the household with a distinction for institutional households [25]; 6) separate comorbidities present throughout the study period (neoplastic disease, liver disease, congestive heart failure, cerebrovascular disease, chronic renal disease, other cardiovascular diseases, pulmonary disease, diabetes mellitus, immunosuppression, neurologic disease, and HIV) based on hospital registrations and medication claims from pharmacies [27]. Comorbidities were classified into larger categories based on the Pneumonia Severity Index (PSI) categories as defined by Fine et al. [28] where possible; 7) the quartile of a year and period of flu season to account for the seasonal and yearly variability in the risk of infection; 8) whether a patient was hospitalised with a complicated LRTI in the past year before observation. Data on smoking status, vaccination status and body mass index were not available in the SN data.

Detailed variable definitions are presented in supplementary table M1. Diagnosis and medication codes used for the definitions of comorbidities and LRTI can be found in the supplementary tables M2 and M3.

### Framework

The variables included were classified as structural or intermediary determinants based on our adaptation of the conceptual framework for action on the social determinants of health from the World Health Organization (WHO) [21] Figure 1). In this framework, structural sociodemographic determinants generate social stratification through structural mechanisms, which may lead to health inequities [21]. Structural sociodemographic determinants are not the direct cause of higher incidence of complicated infections but influence health through intermediary mechanisms such as comorbidities and behaviour. In our study, we recognise that we do not measure all intermediary determinants, and a residual effect of SES and migration background will remain after adjusting for intermediary determinants. However, this reflects clinical practice and population health, in which not all determinants are known, and structural sociodemographic factors can be used as a proxy to identify risk groups.

**Figure 1:**
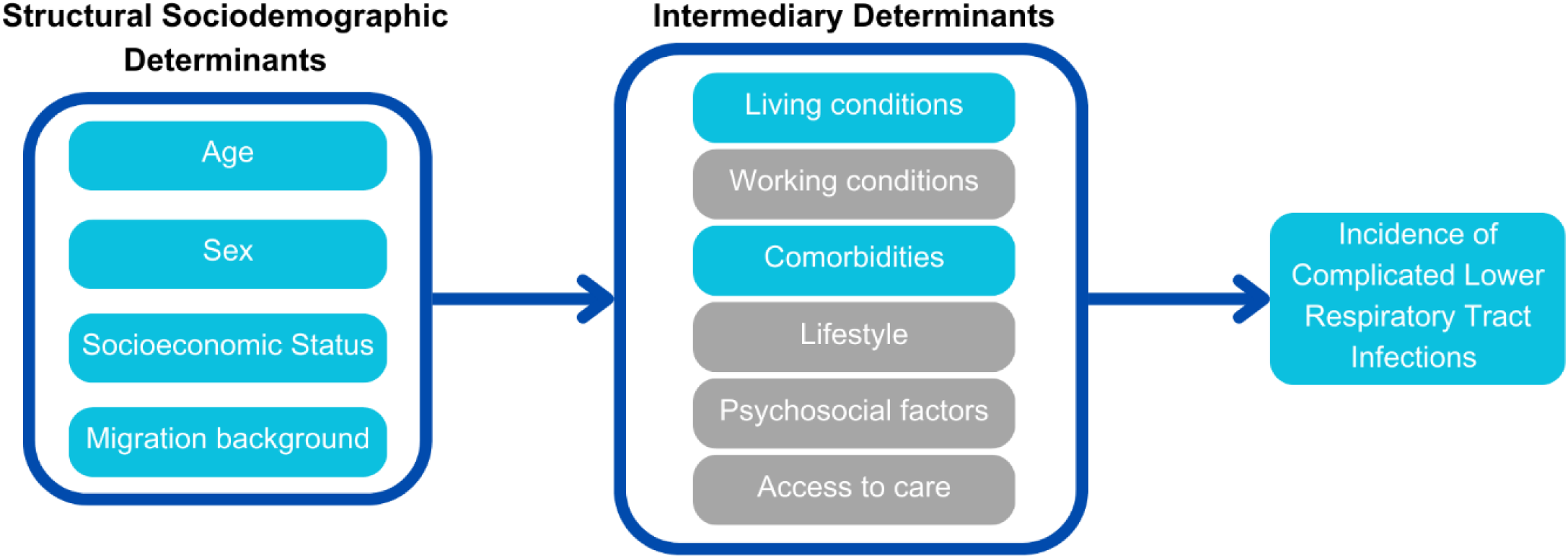
Conceptual framework depicting the role of determinants influencing the incidence of complicated LRTI within this study. Structural sociodemographic determinants influence the incidence of complicated LRTI through various intermediary determinants.

### Statistical Analyses

The characteristics of the overall population and the LRTI cases were described using frequencies and percentages or medians with interquartile range (IQR). The annual complicated LRTI incidence rate was directly standardised for age and sex at birth, using the Dutch population as reference and calculated per 1,000 person-years, overall and by neighbourhood. Neighbourhood SES was defined as the estimated mean of all households within the same postal code. A choropleth heatmap, generated via QGIS, was used to visualise the incidence rates between neighbourhoods in The Hague from 2014 through 2019 and 2020.

Univariable Poisson regression models were used to explore the individual effects of sociodemographic factors on complicated LRTI and LRTI-associated mortality. Age- and SES-stratified analyses were performed to explore the association between migration background and complicated LRTI incidence. Subsequently, determinants were combined into multivariable Poisson regression models. First, multivariable Poisson regression analyses were conducted that included structural sociodemographic determinants (SES, migration background, age, and sex), quartile of a year, flu season, and LRTI hospitalisation in the past year. Thereafter, fully adjusted multivariable Poisson regression analyses were conducted in which the additional intermediary determinants, household composition and comorbidities were added. The possibility of interactions was explored by subsequently adding interaction terms between SES, age, sex, and migration background to the model. Likelihood ratio tests were used to compare models. Time in person-years was used as offset, and clusters on the person level were incorporated to account for the multiple measurements per person for complicated LRTI episodes. The final fully adjusted model included all variables without interaction terms (age, sex, comorbidities, SES, household composition, migration background, quartile of a year, flu season, and hospitalisation for LRTI in the past year). Analyses were performed separately for complicated LRTIs and LRTI–associated mortality as dependent variables. A p-value of <0.05 was considered significant.

Separate analyses were conducted for the 2014–2019 and 2020 periods to compare the determinants of the pre-COVID-19 period with those of the first year of the COVID-19 pandemic. Additionally, individuals aged 80 years and older were excluded from the Poisson regression analyses due to the insufficient number of individuals with a migration background in this group (Supplementary Tables S7 and S8). Furthermore, an exploratory sensitivity analysis was conducted regarding LRTI- associated mortality, accounting for possible deaths of Dutch citizens abroad defaulted to the ICPC-10 code “R99, unknown cause of death” (Supplementary Table S12). Observations were omitted from analyses when data on one of the variables were missing. For none of the variables > 4% was missing. All analyses were performed in Stata (Stata Corp. 2023. Stata Statistical Software: Release 18. College Station, TX: Stata Corp LLC.).

## Results

### Population characteristics

Between 1 January 2014 and 31 December 2019, we identified 5,196 complicated LRTI episodes in 4,821 individuals. From 1 January 2020 to 31 December 2020, 2,303 episodes were identified in 2,285 individuals. The characteristics of the patients with complicated LRTI and the overall population are outlined in Table 1.

**Table 1:** Characteristics of individuals with complicated LRTI episodes and characteristics of the overall population in 2014-2019 and 2020, in The Hague, the Netherlands.

| Variables |  | People with complicated LRTI 2014-2019<sup>a</sup> | Overall Population 2014 – 2019<sup>b</sup> | People with complicated LRTI 2020<sup>a</sup> | Overall Population 2020<sup>b</sup> |
| --- | --- | --- | --- | --- | --- |
| **Number of individuals (number of episodes)** |  | 4,821 (5,196) | 415,945 | 2,285 (2,303) | 434,336 |
| **Sex (% of population)** | **Female** | 2,470 (51.2) | 211,836 (50.9) | 1,023 (44.8) | 220,425 (50.8) |
| **Age in years (% of population)** | **Median (IQR)** | 73.2 (61.1 : 83.8) | 45.2 (31.9 : 59.7) | 72.0 (59.3 : 83.2) | 45.5 (31.9 : 60.0) |
|  | **18-49** | 622 (12.9) | 245,770 (59.1) | 275 (12.0) | 253,638 (58.4) |
|  | **50-59** | 503 (10.4) | 67,949 (16.3) | 319 (14.0) | 71,569 (16.5) |
|  | **60-69** | 896 (18.6) | 51,727 (12.4) | 432 (18.9) | 53,844 (12.4) |
|  | **70-79** | 1,184 (24.6) | 31,498 (7.6) | 508 (22.2) | 35,833 (8.3) |
|  | **80+** | 1,616 (33.5) | 19,001 (4.6) | 751 (32.9) | 19,452 (4.5) |
| **Household SES<sup>c,d</sup> in quintiles (% of the population)** | **1 (highest)** | 576 (12.0) | 78,834 (19.0) | 274 (12.0) | 87,445 (20.1) |
|  | **2** | 488 (10.1) | 70,399 (16.9) | 273 (12.0) | 74,154 (17.1) |
|  | **3** | 692 (14.4) | 72,216 (17.4) | 332 (14.5) | 75,364 (17.4) |
|  | **4** | 1,239 (25.7) | 80,135 (19.3) | 516 (22.6) | 82,656 (19.0) |
|  | **5 (lowest)** | 1,517 (31.5) | 106,703 (25.7) | 766 (33.5) | 106,646 (24.6) |
| **Estimation Comorbidities (% of population)** | **None of listed** | 1,099 (22.8) | 286,895 (69.0) | 589 (25.8) | 303,011 (69.8) |
|  | **Neoplastic Disease** | 997 (20.7) | 57,440 (13.8) | 413 (18.1) | 58,997 (13.6) |
|  | **Liver Disease** | 92 (1.9) | 3,677 (0.9) | 46 (2.0) | 3,780 (0.9) |
|  | **Congestive Heart Failure** | 816 (16.9) | 7,420 (1.8) | 268 (11.7) | 6,462 (1.5) |
|  | **Cerebrovascular Disease** | 492 (10.2) | 9,569 (2.3) | 231 (10.1) | 9,304 (2.1) |
|  | **Renal Disease** | 392 (8.1) | 5,066 (1.2) | 135 (5.9) | 4,549 (1.1) |
|  | **Pulmonary Disease** | 1,349 (28.0) | 16,710 (4.0) | 346 (15.1) | 16,375 (3.8) |
|  | **Dementia** | 203 (4.2) | 3,313 (0.8) | 135 (5.9) | 3,099 (0.7) |
|  | **Neurologic Disease** | 119 (2.5) | 2,753 (0.7) | 60 (2.2) | 2,793 (0.6) |
|  | **Diabetes mellitus** | 1,066 (22.1) | 32,530 (7.8) | 658 (28.8) | 32,014 (7.4) |
|  | **HIV** | 28 (0.6) | 1,187 (0.3) | <10 | 1,227 (0.3) |
|  | **Immunocompromised** | 592 (12.3) | 13,236 (3.2) | 376 (16.5) | 16,875 (3.9) |
|  | **Other Cardiovascular Disease** | 1,040 (21.6) | 23,669 (5.7) | 441 (19.3) | 23,295 (5.4) |
| **Household Composition<sup>e</sup> (% of population)** | **One person household without children** | 1,843 (38.2) | 119,999 (28.9) | 602 (29.6) | 125,596 (28.9) |
|  | **One parent household with children** | 214 (4.4) | 35,734 (8.6) | 90 (5.0) | 37,550 (8.6) |
|  | **Couple without children** | 1,390 (28.8) | 103,286 (24.8) | 597 (30.9) | 106,986 (24.6) |
|  | **Couple with children** | 520 (10.8) | 130,725 (31.4) | 372 (20.7) | 136,301 (31.4) |
|  | **More than two-person adult household without children** | 38 (0.8) | 5,971 (1.4) | 17 (0.9) | 6,733 (1.6) |
|  | **More than two-person adult household with** **children (e.g., multigenerational)** | 93 (1.9) | 9,680 (2.3) | 59 (3.1) | 10,125 (2.3) |
|  | **Institutional household** | 704 (14.6) | 7,522 (1.8) | 541 (23.7) | 8,237 (1.9) |
|  | **Other household type** | 19 (0.4) | 3,028 (0.7) | <10 | 2,769 (0.6) |
| **Migration Background<sup>f</sup>**<br>(% of population) | **The Netherlands** | 3,122 (64.8) | 203,075 (48.8) | 1,113 (48.7) | 198,590 (45.7) |
|  | **Middle and Eastern Europe** | 46 (1.0) | 19,826 (4.8) | 28 (1.2) | 25,950 (6.0) |
|  | **Turkey** | 209 (4.3) | 28,231 (6.8) | 199 (8.7) | 30,115 (6.9) |
|  | **Morocco** | 187 (3.9) | 20,493 (4.9) | 183 (8.0) | 22,095 (5.1) |
|  | **Suriname** | 484 (10.0) | 38,835 (9.3) | 307 (13.4) | 39,385 (9.1) |
|  | **Dutch Caribbean** | 54 (1.1) | 9,866 (2.4) | 40 (1.8) | 10,980 (2.5) |
|  | **Indonesia** | 238 (4.9) | 17,100 (4.1) | 122 (5.3) | 16,451 (3.8) |
|  | **Other Europe** | 277 (5.8) | 33,361 (8.0) | 126 (5.5) | 36,536 (8.4) |
|  | **Other Africa** | 56 (1.2) | 12,618 (3.0) | 61 (2.7) | 14,373 (3.3) |
|  | **Other Asia** | 111 (2.3) | 22,528 (5.4) | 84 (3.7) | 26,262 (6.5) |
|  | **Other America's and Oceania** | 37 (0.8) | 9,999 (2.4) | 22 (1.0) | 11,599 (2.7) |

Abbreviations: SES: socioeconomic status, IQR: interquartile range, SD: standard deviation. Counts of <10 have been suppressed due to potential personal data disclosure. **a.** For 2014–2019: number of patients with complicated LRTI from 1 January 2014 until 31 December 2019. For 2020: number of patients with complicated LRTI from 1 January 2020 until 31 December 2020. **b.** For 2014–2019: population on 1 January 2017. For 2020: population on 1 January 2020. **c.** SES: Socioeconomic status on household level, based on financial prosperity (standardised disposable annual household income + household wealth) provided in percentiles. **d.** 7,658 missing values (1.84%) in the 2014–2019 cohort and 8,071 missing values (1.86%) in the 2020 cohort. **e.** 39 missing variables (0.01%) in the general population cohort in 2020. No missing variables in other cohorts. **f.** 13 missing variables (0.01%) in 2014–2019.

The median age of patients with complicated LRTI was higher in 2014-2019 (73·2 years [IQR: 61·1– 83·8]) and in 2020 (72·0 years [IQR: 59·3–83·2]) compared to the overall population in 2014–2019 (45·2 years [IQR: 31·9–59·7]), and in 2020 (45·5 years [IQR:31·9–60·0]) (Table 1). Sex was approximately equally distributed.

Patients with complicated LRTI were more likely to have a comorbid medical condition in both 2014-2019 (77%) and 2020 (74%) compared to the overall population (31% and 30%, respectively; Table 1). Furthermore, individuals in the fourth and fifth SES quintiles, corresponding to lower SES, were overrepresented in the complicated LRTI patients compared to the overall population in both periods (Table 1). Moreover, complicated LRTI patients were more likely to have no migration background compared to the overall population, particularly in 2014-2019 (Table 1). In 2020, complicated LRTI patients were more likely to be Turkish, Moroccan, Surinamese, and Indonesian compared to the overall population (Table 1).

### Geographic Incidence

The estimated crude annual complicated LRTI incidence rates were 2.08 (95% CI: 2.02–2.13) per 1,000 person-years from 2014–2019 and 5.26 (95% CI: 5.05–5.48) per 1,000 person-years in 2020. Higher age- and sex-standardised complicated LRTI incidence rates (SIR) were observed in more socioeconomically deprived neighbourhoods across both periods (Figure 2).

**Figure 2:**
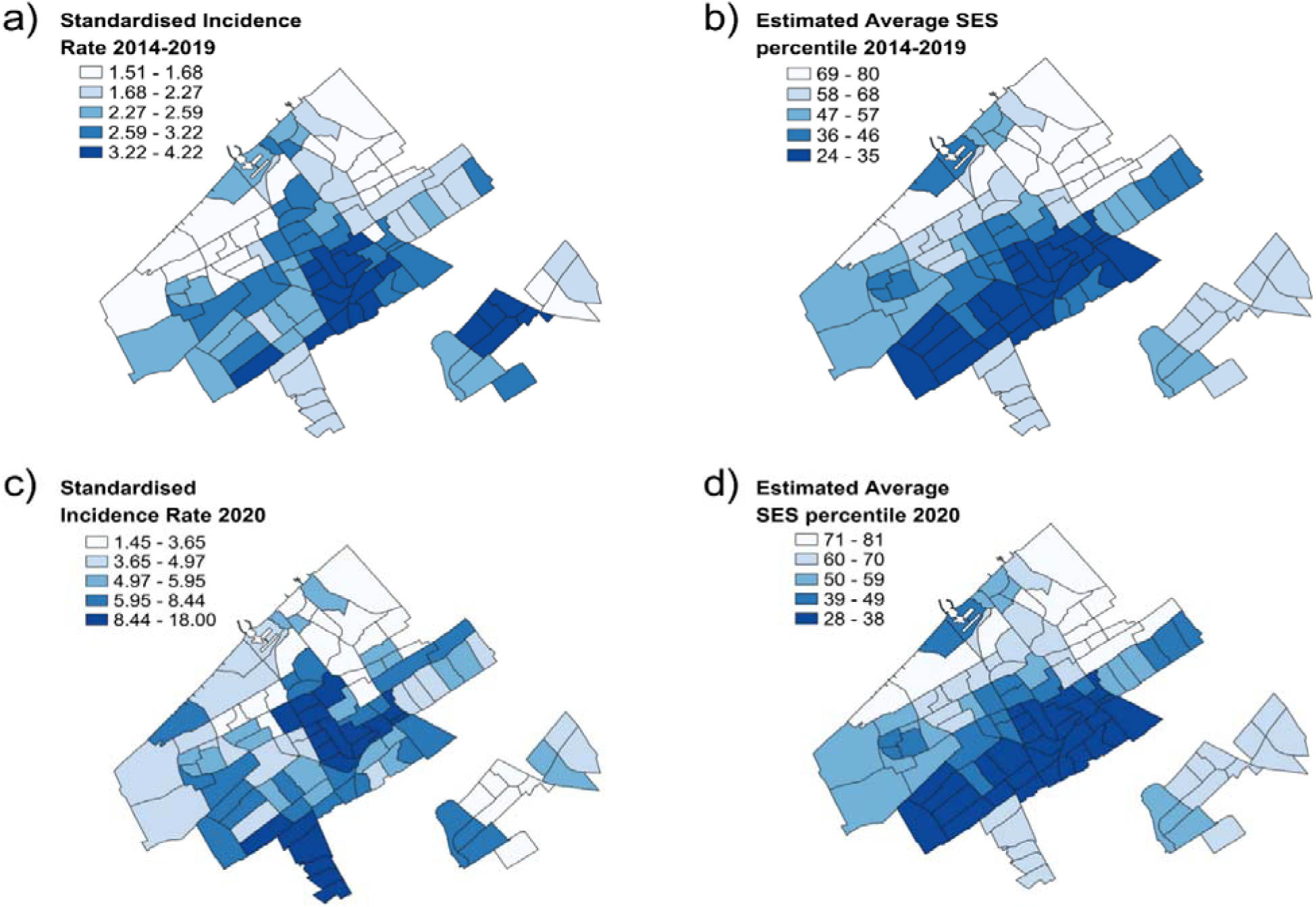
Choropleth heatmaps of the age- and sex Standardised Incidence Rates (SIR) of complicated LRTI, and average SES percentiles per neighbourhood in The Hague, 2014–2019 and 2020. Rates per 1,000 person-years standardised to the standard Dutch population. Heatmaps of the age- and sex standardised incidence rates (SIR) per 1,000 person-years of complicated LRTI per neighbourhood in The Hague for 2014–2019 **a)** and 2020 **c)**. For comparison, the estimated average SES percentile per neighbourhood in The Hague for 2014–2019 and 2020 are shown in **b)** and **d),** respectively. Neighbourhoods are based on the 4-digit postal code. A darker colour indicates a more socioeconomic deprived neighbourhood with lower average SES.

In 2014–2019, the neighbourhood with the lowest average SES had a SIR of 3.22 (95% CI: 2.72– 3.73) per 1,000 person-years, whereas the SIR of the neighbourhood with the highest average SES was 1.65 (95% CI: 1.19–2.12) per 1,000 person-years.

Similar disparities were observed in 2020, where the neighbourhood with the lowest average SES had a SIR of 7.15 (95% CI: 5.44–8.87) per 1,000 person-years, compared with 4.06 (95% CI: 2.30–5.82) per 1,000 person-years in the neighbourhood with the highest average SES.

### Sociodemographic determinants associated with complicated LRTI incidence and LRTI-associated mortality

We stratified crude complicated LRTI incidence rates by age and SES based on migration background for 2014-2019 (Supplementary Tables S7 and S9) and 2020 (Supplementary Tables S8 and S10). Trends in incidence rates were similar across people with and without migration background when stratified by age and SES.

In 2014-2019, lower SES was consistently associated with increased adjusted incidence rate ratios (aIRR) of complicated LRTI (Table 2A). This association was partially mediated by comorbidities and household composition, as reflected by a reduced aIRR of the lowest SES quintile in the full model (aIRR = 2.26 [95%CI: 1.98 – 2.57]) compared to the structural determinants model (aIRR = 3·55 [95%CI: 3·12 – 4·05]); Table 2A). Furthermore, most migration background populations were associated with lower complicated LRTI incidence, except for Turkish, Moroccan and Surinamese groups (Table 2A).

**Table 2A and 2B.** Univariable, multivariable with structural determinants, and full multivariable Poisson regression analyses of determinants associated with complicated LRTI incidence, between the ages of 18 and 80 years, in 2014–2019 (A) and 2020 (B).

| 2A |  |  |  | 2B |  |  |  |
| --- | --- | --- | --- | --- | --- | --- | --- |
| Independent Variables | Jan 2014 – Dec 2019 |  |  | Independent Variables | Jan 2020 – Dec 2020 |  |  |
|  | IRR<sup>a</sup> (95% CI) | aIRR<sup>b</sup> (95% CI)<br>[structural determinants] | aIRR<sup>c</sup> (95% CI)<br>[all determinants] |  | IRR<sup>a</sup> (95% CI) | aIRR<sup>b</sup> (95% CI)<br>[structural determinants] | aIRR<sup>c</sup> (95% CI)<br>[all determinants] |
| **Sex** |  |  |  | **Sex** |  |  |  |
| Male | Ref | Ref | Ref | Male | Ref | Ref | Ref |
| Female | 0.92 (0.85 – 0.98) | 0.85 (0.79 – 0.92) | 0.84 (0.78 – 0.90) | Female | 0.66 (0.59 – 0.73) | 0.64 (0.57 – 0.70) | 0.64 (0.57 – 0.71) |
| **Age<sup>d</sup>** |  |  |  | **Age<sup>d</sup>** |  |  |  |
| 18–49 | Ref | Ref | Ref | 18–49 | Ref | Ref | Ref |
| 50 – 59 | 2.99 (2.65 – 3.37) | 2.97 (2.63 – 3.37) | 2.20 (1.93 – 2.49) | 50 – 59 | 4.08 (3.47 – 4.79) | 4.36 (3.70 – 5.13) | 3.36 (2.84 – 3.98) |
| 60 – 69 | 7.27 (6.54 – 8.07) | 7.18 (6.44 – 8.00) | 4.28 (3.79 – 4.83) | 60 – 69 | 7.50 (6.45 – 8.73) | 8.42 (7.21 – 9.84) | 5.45 (4.58 – 6.48) |
| 70 – 80 | 15.82 (14.33 – 17.46) | 14.76 (13.28 – 16.41) | 7.28 (6.43 – 8.25) | 70 – 80 | 12.96 (11.19 – 15.00) | 15.61 (13.38 – 18.21) | 8.20 (6.82 – 9.86) |
| **Household SES in quintiles** |  |  |  | **Household SES in quintiles** |  |  |  |
| 1 (highest) | Ref | Ref | Ref | 1 (highest) | Ref | Ref | Ref |
| 2 | 1.07 (0.92 – 1.24) | 1.35 (1.16 – 1.56) | 1.18 (1.02 – 1.37) | 2 | 1.24 (1.02 – 1.52) | 1.38 (1.13 – 1.68) | 1.26 (1.03 – 1.54) |
| 3 | 1.41 (1.23 – 1.62) | 1.78 (1.55 – 2.05) | 1.47 (1.28 – 1.69) | 3 | 1.50 (1.24 – 1.81) | 1.60 (1.32 – 1.93) | 1.39 (1.15 – 1.69) |
| 4 | 1.99 (1.75 – 2.27) | 2.15 (1.89 – 2.45) | 1.59 (1.39 – 1.81) | 4 | 1.78 (1.49 – 2.13) | 1.69 (1.41 – 2.04) | 1.38 (1.14 – 1.66) |
| 5 (lowest) | 2.48 (2.20 – 2.80) | 3.47 (3.06 – 3.94) | 2.26 (1.98 – 2.57) | 5 (lowest) | 2.36 (2.00 – 2.78) | 1.99 (1.66 – 2.38) | 1.47 (1.21 – 1.77) |
| **Migration Background** |  |  |  | **Migration Background** |  |  |  |
| The Netherlands | Ref | Ref | Ref | The Netherlands | Ref | Ref | Ref |
| Middle & Eastern Europe | 0.20 (0.15 – 0.28) | 0.42 (0.30 – 0.58) | 0.51 (0.37 – 0.72) | Middle & Eastern Europe | 0.31 (0.21 – 0.46) | 0.83 (0.56 – 1.25) | 0.99 (0.66 – 1.48) |
| Turkey | 0.69 (0.59 – 0.81) | 1.00 (0.85 – 1.17) | 0.99 (0.84 – 1.16) | Turkey | 1.96 (1.65 – 2.31) | 3.36 (2.81 – 4.02) | 2.83 (2.35 – 3.40) |
| Morocco | 0.88 (0.74 – 1.04) | 1.00 (0.84 – 1.18) | 1.10 (0.93 – 1.30) | Morocco | 2.46 (2.07 – 2.93) | 3.53 (2.93 – 4.25) | 3.19 (2.63 – 3.89) |
| Suriname | 1.12 (1.00 – 1.24) | 1.23 (1.10 – 1.38) | 1.08 (0.96 – 1.21) | Suriname | 2.19 (1.89 – 2.54) | 2.60 (2.23 – 3.02) | 2.00 (1.71 – 2.34) |
| Dutch Caribbean | 0.49 (0.37 – 0.66) | 0.64 (0.47 – 0.86) | 0.72 (0.53 – 0.97) | Dutch Caribbean | 1.04 (0.74 – 1.45) | 1.68 (1.19 – 2.37) | 1.69 (1.19 – 2.40) |
| Indonesia | 0.76 (0.63 – 0.92) | 0.72 (0.60 – 0.87) | 0.78 (0.65 – 0.94) | Indonesia | 1.57 (1.23 – 2.01) | 1.35 (1.05 – 1.73) | 1.42 (1.11 – 1.83) |
| Other Europe | 0.56 (0.48 – 0.66) | 0.75 (0.64 – 0.87) | 0.81 (0.69 – 0.94) | Other Europe | 0.65 (0.51 – 0.84) | 1.04 (0.81 – 1.33) | 1.12 (0.87 – 1.44) |
| Other Africa | 0.39 (0.30 – 0.51) | 0.57 (0.43 – 0.76) | 0.62 (0.47 – 0.83) | Other Africa | 1.27 (0.97 – 1.67) | 2.15 (1.63 – 2.83) | 2.07 (1.57 – 2.74) |
| Other Asia | 0.42 (0.33 – 0.52) | 0.59 (0.47 – 0.74) | 0.68 (0.54 – 0.86) | Other Asia | 0.77 (0.60 – 0.99) | 1.31 (1.01 – 1.71) | 1.34 (1.03 – 1.75) |
| Other America's and Oceania | 0.34 (0.24 – 0.49) | 0.61 (0.42 – 0.88) | 0.70 (0.49 – 1.01) | Other America's and Oceania | 0.57 (0.37 – 0.88) | 1.22 (0.79 – 1.88) | 1.35 (0.87 – 2.09) |

Tables for all univariable Poisson regression models and multivariable Poisson regression models with all prespecified determinants included can be found in Supplementary Tables S3–S6. **a.** Univariable unadjusted incidence rate ratio (IRR) **b.** Adjusted incidence rate ratio (aIRR) for a multivariable model with structural determinants SES, Migration background, age, and sex, and quartile of a year, flu season, and LRTI hospitalisation in the past year. **c.** Adjusted incidence rate ratio for the full multivariable model with SES, migration background, age, sex, household composition, comorbidities, flu season, quartile of a year, and LRTI hospitalisation in the past year **d.** Those above 80 years of age were excluded from the regression analyses due to the small number of individuals over 80 years of age in the migration background groups.

In 2020, lower SES remained consistently associated with increased aIRR of complicated LRTI (Table 2B). Similarly to 2014-2019, this association was evident in the structural determinants model (aIRR = 1.99 [95%CI: 1.66 – 2.38]) and remained in the full model (aIRR = 1.47 [95%CI: 1.21 – 1.77]) (Table 2B). In contrast to 2014-2019, migration background was associated with higher complicated LRTI incidence rates. When adjusted for all prespecified determinants, particularly those with Turkish (aIRR = 2.83 [95% CI: 2.35 – 3.40]), Moroccan (aIRR = 3.19 [95%CI: 2.63 – 3.89]), and Surinamese (aIRR = 2.00 [95%CI: 1.71 – 2.34]) backgrounds showed elevated aIRR compared to those without a migration background (Table 2B).

LRTI-associated mortality analyses showed a similar association with the lowest SES quintile in 2014-2019 (aIRR = 3.49 [95%CI: 2.28 – 5.34]; Supplementary Table S5). However, contrary to the complicated LRTI-incidence analyses, a Turkish background was associated with lower LRTI-associated mortality incidence (aIRR = 0.42 [95%CI: 0.20 – 0.86] in the full model (Supplementary Table S5). In the sensitivity analysis incorporating the ICD-10 code “R99, unknown cause of death” (which is used for inhabitants that decease abroad) in our LRTI-associated mortality outcome, being of Turkish descent was no longer significantly associated with lower mortality rates as the aIRR increased to 0.80 (95% CI: 0.48–1.34) (Supplementary Table S12). The aIRR of those with a Moroccan and Surinamese background also increased compared with the initial multivariable analysis regarding LRTI-associated mortality (Supplementary Table S12).

In 2020, patterns regarding LRTI-associated mortality incidence rates mirrored the trends seen for complicated LRTI incidence, where the lowest SES quintile showed an aIRR of 2.59 (95% CI: 1.61 – 4.16) in the full model (Supplementary Table S6).

Tables presenting all prespecified determinants for univariable and multivariable Poisson regression models can be found in Supplementary Tables S3-S6.

## Discussion

This population-based cohort study identified substantial health disparities in complicated LRTI incidence in The Hague, The Netherlands. Complicated LRTI cases were clustered in more socioeconomic deprived neighbourhoods before and during the COVID-19 pandemic. We found consistent strong associations between lower SES and higher incidence and mortality of complicated LRTI. These associations were only partly mediated by comorbidities and household composition. Contrary to the pre-COVID-19 period, complicated LRTI was strongly associated with migration background during the first year of the COVID-19 pandemic, particularly among Turkish, Moroccan, and Surinamese population groups - the largest populations with a migration background in The Netherlands.

Previous studies on CAP and influenza in the United States and the United Kingdom similarly demonstrated health disparities linked to SES and area deprivation regarding complicated LRTI [11–14]. Our findings extend this evidence to The Netherlands, a country with a healthcare system resembling those in other West European countries. Furthermore, we could evaluate the role of comorbidity and household composition as intermediary factors. We found that even after adjusting for these factors, SES remains strongly associated with complicated outcomes, underscoring its role as a proxy for a complex cluster of risk factors. Other mediating factors include housing quality, environmental pollution, working conditions, access to care, vaccination uptake, health literacy, psychological factors, and lifestyle factors, including smoking and obesity [21, 29–32]. In socioeconomically deprived neighbourhoods, these factors converge [33, 34], aligning with our findings of higher complicated LRTI incidence in these areas. Co-designed community outreach programs targeting lower SES neighbourhoods could help enhance health education and development of targeted vaccination initiatives, which could serve as initial steps toward mitigating health inequities in the population regarding complicated LRTI.

Contrary to our findings, a study from the United States demonstrated ethnic disparities in influenza-associated hospitalisation and mortality before the COVID-19 pandemic [9]. We found that, on average, people with a migration background had lower SES than the general population, suggesting that studies not accounting for SES may have attributed SES-driven disparities to migratory background. In our 2014-2019 data, an initially observed lower LRTI-associated mortality rate in individuals of Turkish background disappeared after adjusting for deaths occurring abroad, as shown in our sensitivity analysis. This suggests that the apparent protective effect of migration background may partly reflect individuals spending significant time abroad, where LRTI-related deaths may occur unreported. Therefore, the true effect of migration background in 2014-2019 may have been underestimated. Additionally, younger migrant populations, such as individuals born in Eastern Europe, are often working migrants and generally healthier than the average population, a phenomenon known as the “healthy immigrant effect” [35], likely contributing to their lower complication rates.

The discrepancy regarding the effect of migration background between 2014-2019 and 2020 is likely due to the pandemic’s distinct societal and pathophysiological characteristics. The health disparities regarding migration background observed in our 2020 cohort align with findings from other Dutch COVID-19 studies [17, 19]. Our study contributes to this body of evidence by incorporating household SES, comorbidities, and household composition as covariates. Notably, migratory background remained a significant factor after adjusting for comorbidities, indicating that the theory presented by Collard et al. [17], stating that the high burden of pre-existing comorbidities potentially drives the higher risk of hospital admission, is at least not the only underlying mechanism. Obesity, which disproportionally affects populations with a migration background [36], likely plays a role, as it increases LRTI disease severity, particularly of COVID-19 [37]. Furthermore, higher infection rates, driven by working and living conditions and limited adherence to preventive measures, may also contribute [16]. Low health literacy, language barriers, and inadequate access to culturally appropriate information likely hinder symptom recognition, adequate prevention methods and timely care [16, 17, 38], heightening the likelihood of complications. Moreover, higher antibiotic prescription rates among migrant populations [39], while potentially offering some protection before the pandemic, did not improve outcomes during COVID-19 and may have worsened disease severity [40].

Our study had several strengths. A major strength is the size and diversity of the study population, entailing all adult residents of The Hague over six years. More than half of the population of The Hague has a migration background, and The Hague has the highest income-based segregation in The Netherlands. Furthermore, by analysing determinants of LRTI complications before and after the onset of the COVID-19 pandemic, we could observe changes in the effect of determinants. Additionally, including SES, migration background, comorbidities, and household composition allowed for a nuanced understanding of the relative associations of these structural and intermediary factors. However, this study also has some limitations. First, data on other intermediary determinants were unavailable. More research is needed to understand the contribution of these factors better. Nonetheless, structural determinants can serve as proxies for unmeasured intermediary determinants when designing population health strategies and treatment guidelines. Second, reliance on insurance claims data may introduce misclassification bias. However, our use of diagnostic imaging to define outcomes minimises this risk. Furthermore, our reported comorbidity rates are consistent with those in prior prospective LRTI studies. Lastly, the distinct population of The Hague might limit the generalisability of results. However, as our findings align with those found in studies from the United States and the United Kingdom, we expect our findings to provide relevant insights for other major cities with diverse populations and income-based segregation.

In conclusion, our study shows significant health disparities in complicated LRTI in The Hague with socioeconomic area deprivation, lower SES and migration background strongly linked with complicated infections. While migration background was not linked to LRTI complications pre-pandemic, its significant association with adverse outcomes during the COVID-19 pandemic underscores the differential impact of risk factors in a pandemic context. These results emphasise the need for further research into other mediating factors and targeted interventions, such as incorporating SES as a risk factor in clinical decision-making and focusing population- and public health strategies on more deprived areas.

## Data availability statement

The dataset cannot be shared directly due to the current Dutch legislation for data protection. Under certain conditions, the dataset and additional microdata are accessible for statistical and scientific research and must be directly requested from Statistics Netherlands (.). The Stata scripts used will be disclosed through a repository on GitHub: https://github.com/elan-dcc/VanDokkum25.

## Ethics statements

### Patient and public involvement

Patients and/or the public were not involved in the design, conduct, reporting, or dissemination plans of this research.

### Patient consent for publication

Not applicable

### Ethics approval

The appropriate approval that the study was not subject to the Medical Examination Act was granted by the Medical Ethical Committee of the Leiden University Medical Center (LUMC) under reference number 24-3031. The study protocol was approved by the Scientific Review Board of the Department of Public Health and Primary Care of the LUMC in 2024.

## Footnotes

### Author contributions

All authors contributed equally to the conceptualisation of the study. Study design and methodology: HB CvN EDvD SlC. Literature search: EDvD with contributions from HB and CvN. Data curation and data analysis: EDvD and HB with substantial contributions from SlC. Data interpretation: EDvD HB and CvN with substantial contributions from all authors. Visualisation of results: EDvD HB and CvN with contributions of all authors. EDvD wrote the first draft of the manuscript with inputs from HB and CvN. All authors reviewed and revised drafts of the manuscript and approved the final version. EDvD and HB have full access to and have verified all the study data provided for the analysis.

