## Supplementary Material for "Health Disparities in Complicated Lower Respiratory Tract Infections: a population-based cohort study in The Hague, The Netherlands"

**M-Table**

**Table M1** – Definitions of determinants used in the study

**Table M2** – DTC-codes used for definition of LRTI and comorbidities

**Table M3** – Drugs by ATC4-codes, used to help define Diabetes Mellitus and Immunosuppression.

**S-Tables**

**Table S1** – Crude, and age- and sex standardised complicated LRTI incidence rates per 1,000 person-years by neighbourhood in The Hague, over 18 years of age in 2014-2019 and 2020

**Table S2** – Estimated average household SES percentile by neighbourhood in The Hague in 2014-2019 and 2020

**Table S3** – Univariable, multivariable with structural determinants, and full multivariable Poisson regression analyses of determinants associated with complicated LRTI incidence, between the ages of 18 and 80, in 2014-2019.

**Table S4** – Univariable, multivariable with structural determinants, and full multivariable Poisson regression analyses of determinants associated with complicated LRTI incidence, between the ages of 18 and 80, in 2020.

**Table S5** – Univariable, multivariable with structural determinants, and full multivariable Poisson regression analyses of determinants associated with LRTI-associated mortality, between the ages of 18 and 80, in 2014-2019.

**Table S6** – Univariable, multivariable with structural determinants, and full multivariable Poisson regression analyses of determinants associated with LRTI-associated mortality, between the ages of 18 and 80, in 2020.

**Table S7** – Age group specific, crude complicated LRTI incidence, LRTI-associated mortality, and overall mortality rates per 1,000 person-years by migration background, over 18 years of age in 2014-2019.

**Table S8** – Age group specific, crude complicated LRTI incidence, LRTI-associated mortality, and overall mortality rates per 1,000 person-years by migration background, over 18 years of age in 2020.

**Table S9** – SES-quintile specific, crude complicated LRTI incidence, LRTI-associated mortality, and overall mortality rates per 1,000 person-years by migration background, over 18 years of age in 2014-2019.

**Table S10** – SES-quintile specific, crude complicated LRTI incidence, LRTI-associated mortality, and overall mortality rates per 1,000 person-years by migration background, over 18 years of age in 2020.

**Table S11** – SES-quintile specific, crude complicated LRTI incidence rates per 1,000 person-years, by flu-season, over 18 years of age.

**Table S12** – Fully adjusted multivariable Poisson regression sensitivity analysis regarding LRTI-associated mortality, which includes deaths occurring abroad in the composite outcome measure, between the ages of 18 and 80, in 2014-2019.

**M-References**

**Table M1 – Definitions of determinants used in the study**

| **Determinants** | **Definition** |
| --- | --- |
| **Complicated Lower Respiratory Tract Infection** | In the Dutch healthcare system, hospital care payment claims are based on Diagnosis Treatment Combinations (DTCs). Every patient receiving hospital care is assigned a specific DTC claim corresponding to the diagnosis and intervention the patient has underwent. Through these DTCs, information could be obtained on which department(s) a patient visited at, the subsequent diagnosis, what interventions or diagnostic screening tools were performed, and whether a patient was admitted. Through these DTC claims we defined a complicated LRTIcase and comorbidities.  Our primary outcome, a complicated LRTI case, was defined as a patient with a pneumonia or influenza-like illness (ILI) diagnosis between January 1^st^, 2014, and December 31^st^, 2020, who visited the emergency department (ED), received chest imaging (X-ray or Computed Tomography) and was subsequently admitted to the hospital or died within 14 days from the ED visit. Furthermore, to focus solely on community-acquired cases, patients discharged from a hospital within 30 days before the ED visit were excluded to avoid including hospital-acquired cases. Those who died with LRTI as a registered cause of mortality, irrespective of an ED visit, were also included, as this was regarded as a complicated LRTI. |
| **Age** | Age at observation date |
| **Sex** | Sex assigned at birth |
| **City neighbourhoods** | Neighbourhoods of the city of The Hague were determined based on a four-digit postal code of current residence. The first two digits of the postal code indicate a city and region, and the second two digits indicate a range of house numbers in the same neighbourhood [1]. |
| **Household Socioeconomic Status (SES)** | SES was defined at the household level as financial prosperity using a composite measure of the household’s standardised annual disposable income and wealth [2]. SN provided household financial prosperity in percentiles (ranging from 1–100) within the entire Dutch population. Household wealth encompasses the total value of financial assets, extracting any outstanding liabilities [2]. To ensure accuracy, household financial prosperity from the previous year rather than the current year was used, accounting for potential major reductions in annual income due to severe illness or death. Additionally, if SES data were missing in the year prior to the observation, the available SES data closest to the particular year were used. Household SES for students and individuals living in institutional households was approximated through the available SES data closest to the particular year, or classified as missing (3%) if no data were available throughout the study period.  SES was divided into quintiles for analysis ranked from the wealthiest 20% of the Dutch population (Q1) to the 20% poorest of the Dutch population (Q5). |
| **Migration background** | The term ‘migration background’ was used for foreign-born individuals and individuals born in the Netherlands with one or two parents born abroad. Migration background was determined by an individual’s country of birth when born abroad or the parent’s country of birth when the individual was born in The Netherlands [2]. If both parents were born abroad, migration background was determined based on the mother’s country of birth [2]. When an individual and their mother were born in the Netherlands, migration background was defined by the country of birth of the non-Dutch father [2]. |
| **Household composition** | Household composition was defined based on the number of adults and children living in the household. An institutional household was defined as a single accommodation where two or more people reside and whose housing and daily needs are provided professionally, such as nursing homes [2]. |
| **Comorbidities** | Comorbidities were identified through additional DTC registrations and medication claims. These DTCs were classified into larger categories based on the Pneumonia Severity Index (PSI) categories as defined by Fine et al.: neoplastic disease, liver disease, congestive heart failure, cerebrovascular disease, and chronic renal disease [3]. In addition, the presence of other cardiovascular diseases, pulmonary disease, diabetes mellitus, immunosuppression, neurologic disease, and HIV infection was assessed though DTC-claims. Chronic comorbidities, for which DTCs started in 2014–2020, were assumed to be present throughout the study period, as it is highly likely that the disease was already present before the first hospital visit.  By solely assessing DTCs, information on relevant comorbidities diagnosed or treated by a general practitioner could be missed. Therefore, the use of immunosuppressants and diabetes medication was identified through medication claims of pharmacies based on the Anatomical Therapeutic Chemical (ATC) codes [4]. Patients were considered immunocompromised if at least one immunosuppressant was used in the year of observation or if a DTC was opened that suggested impaired immunity in the year of observation. The diabetes group encompassed patients using diabetes medication, having a diabetes DTC, or both at any point during the study period.  **All DTC-codes and ATC-codes that were used to define comorbidities can be found in Supplementary Table M2 and M3.** |
| **Quartile of a year** | The quartile of a year and the period of flu season were included as covariates to account for the seasonal and yearly variability in the risk of infection  Quartile 1 = 1^st^ of January – 31^st^ of March  Quartile 2 = 1^st^ of April – 30^th^ of June  Quartile 3 = 1^st^ of July – 30^th^ of September  Quartile 4 = 1^st^ of October – 31^st^ of December |
| **Period of flu season** | The quartile of a year and the period of flu season were included as covariates to account for the seasonal and yearly variability in the risk of infection.  Flu season 13/14 = First two quartiles of 2014  Flu season 14/15 = Quartile 3 & 4 of 2014 and quartile 1 & 2 of 2015  Flu season 15/16 = Quartile 3 & 4 of 2015 and quartile 1 & 2 of 2016  Flu season 16/17 = Quartile 3 & 4 of 2016 and quartile 1 & 2 of 2017  Flu season 17/18 = Quartile 3 & 4 of 2017 and quartile 1 & 2 of 2018  Flu season 18/19 = Quartile 3 & 4 of 2018 and quartile 1 & 2 of 2019  Flu season 19/20 = Quartile 3 & 4 of 2019 |
| **Hospitalisation with LRTI in the past year** | As a previous complicated LRTI might also increase the risk of a future complicated LRTI, a covariate was included on whether a patient was hospitalised with a complicated LRTI in the past year. This was defined as an individual who had been hospitalised with a LRTI within the 365 days prior to the observation. |

**Table M2 – DTC-codes used for definition of LRTI and comorbidities**

| **Definition** | **DTC-codes** |
| --- | --- |
| **Lower respiratory tract infection** | 0313#401, 0313#409, 0313#469, 0313#601, 0313#402, 0322#1401, 0322#4, 0322#1405, 0322#5663 |
| **Neoplastic disease [3]** | 0302#21, 0302#60, 0302#61, 0302#62, 0302#63, 0302#64, 0302#65, 0302#66, 0302#67, 0302#68, 0302#69, 0302#72, 0302#84, 0303#303, 0303#306, 0303#318, 0303#319, 0303#331, 0303#332, 0303#333, 0303#334, 0303#335, 0303#346, 0303#347, 0303#349, 0303#350, 0303#352, 0303#353, 0303#357, 0303#358, 0303#360, 0303#363, 0303#367, 0303#370, 0305#1110, 0305#1140, 0305#1150, 0306#10, 0306#16, 0306#20, 0306#30, 0306#40, 0306#45, 0306#48, 0306#50, 0306#60, 0306#69, 0306#70, 0306#78, 0306#84, 0306#92, 0307#M11, 0307#M12 0307#M13 0307#M14, 0307#M15, 0307#M16, 0307#M99, 0313#214, 0313#264, 0313#621, 0313#622, 0313#623, 0313#624, 0313#629, 0313#751, 0313#752, 0313#753, 0313#754, 0313#756, 0313#757, 0313#761, 0313#771, 0313#801, 0313#802, 0313#811, 0313#821, 0313#822, 0313#823, 0313#831, 0313#832, 0313#833, 0313#834, 0313#839, 0313#841, 0313#842, 0313#843, 0313#899, 0313#904, 0313#914, 0313#964, 0313#979, 0318#307, 0318#407, 0318#408, 0318#610, 0318#712, 0318#735, 0322#1303, 0322#1304, 0322#1305, 0322#1306, 0322#1308, 0330#202, 0330#203, 0330#213, 0330#223, 0330#233, 0330#242, 0330#243. |
| **Liver disease [3]** | 0313#463, 0313#941 0313#942, 0313#943, 0313#944, 0313#945, 0313#946, 0318#701, 0318#705, 0318#707, 0318#708, 0318#709, 0318#713, 0318#718. |
| **Congestive heart failure [3]** | 0313#107, 0320#301, 0320#302, 0335#262. |
| **Cerebrovascular disease [3]** | 0313#121, 0330#1102, 0330#1111, 0330#1112, 0335#263. |
| **Chronic renal disease [3]** | 0313#324, 0313#325, 0313#331, 0313#332, 0313#336, 0313#339. |
| **Pulmonary disease (other)** | 0303#309, 0313#601, 0322#1201, 0322#1241, 0322#1403, 0335#272. |
| **Dementia** | 0313#91, 0330#401. |
| **Neurologic disease** | 0330#501, 0330#522, 0330#531, 0330#911, 0330#999, 0335#252. |
| **Diabetes Mellitus** | 0313#221, 0313#222, 0313#223, 0318#902, 0335#222 |
| **HIV** | 0313#461, 0313#462. |
| **Immunocompromised** | 0303#551, 0303#553, 0303#554, 0303#555, 0303#557, 0303#559, 0303#560, 0303#561, 0303#562, 0303#563, 0313#70, 0313#72, 0313#73, 0313#74, 0313#76, 0313#78, 0313#79, 0313#81, 0313#82, 0313#83, 0318#761 0318#763 0318#764, 0318#766, 0318#767, 0318#768, 0328#2910, 0328#2920, 0328#2930, 0303#325, 0303#326, 0305#1394, 0313#501, 0313#503, 0313#512, 0313#515, 0313#521, 0313#522, 0313#523, 0313#524, 0313#525, 0313#526, 0313#527, 0313#922, 0313#923, 0318#601, 0318#602, 0324#101, 0324#102, 0324#114, 0324#201, 0324#202, 0324#301, 0324#302, 0324#304, 0324#305, 0324#306, 0324#307, 0324#311, 0324#312, 0324#313, 0324#315, 0324#316, 0324#317, 0324#318, 0324#319. |
| **Other cardiovascular disease** | 0303#412 0303#418, 0303#419, 0303#420, 0313#124, 0313#133, 0320#202 0320#203, 0320#204, 0320#205, 0320#601, 0320#801, 0320#802, 0320#803, 0320#804, 0328#2220, 0328#2320, 0328#2400, 0328#2415, 0328#2425, 0328#2470, 0328#2550, 0328#2555, 0328#2560, 0328#2570, 0328#2585, 0328#2630, 0328#2635, 0328#2640, 0328#2645, 0328#2650, 0328#2655, 0328#2665, 0328#2720, 0328#2740, 0328#2770, 0328#2785, 0328#2940, 0328#3210, 0328#3310 |

**Table M3 – Drugs by ATC4-code used to define diabetes mellitus and immunosuppression**

| **ATC4-code [4]** | **Name** |
| --- | --- |
| **Diabetes Mellitus Medication** | |
| A10A | Insulins and analogues |
| A10B | Blood glucose lowering drugs, excluding insulins |
| **Immunosuppressives** | |
| L04A | Immunosuppressive drugs |
| L01B | Antimetabolites |
| L01X | Other antineoplastic agents |
| H02A | Corticosteroids for systemic use, plain |

**Table S1 – Crude, and age- and sex standardised complicated LRTI incidence rates per 1,000 person-years by neighbourhood in The Hague, over 18 years of age in 2014-2019 and 2020.**

| **Postal code** | **Crude Incidence Rate (IR) per 1,000 person-years**  **(95% CI)**  **2014-2019** | **Standardised Incidence Rate* (SIR) per 1,000 person-years (95% CI)**  **2014-2019** | **Crude Incidence Rate (IR) per 1,000 person-years (95% CI)**  **2020** | **Standardised Incidence Rate* (SIR) per 1,000 person-years (95% CI)**  **2020** |
| --- | --- | --- | --- | --- |
| 2492 | 1.46 (1.18 - 1.81) | 2.00 (1.63 – 2.37) | 3.96 (2.88 - 5.45) | 4.39 (3.06 – 5.72) |
| 2493 | 1.12 (0.80 - 1.55) | 1.60 (1.16 – 2.04) | 2.71 (1.63 - 4.50) | 5.95 (3.92 – 7.98) |
| 2496 | 1.45 (1.15 - 1.84) | 3.25 (2.74 – 3.77) | 2.31 (1.48 - 3.63) | 3.25 (2.02 – 4.48) |
| 2497 | 1.50 (1.18 - 1.90) | 2.33 (1.88 – 2.77) | 5.23 (3.85 - 7.11) | 6.74 (4.92 – 8.55) |
| 2498 | 0.96 (0.63 - 1.46) | 2.65 (1.98 – 3.31) | 1.00 (0.38 - 2.67) | 1.45 (0.27 – 2.63) |
| 2511 | 0.88 (0.57 - 1.36) | 1.63 (1.10 – 2.15) | 3.95 (2.42 - 6.44) | 6.06 (3.66 - 8.45) |
| 2512 | 2.06 (1.75 - 2.42) | 3.64 (3.19 – 4.08) | 4.41 (3.37 - 5.77) | 6.38 (4.95 – 7.81) |
| 2513 | 1.74 (1.37 - 2.21) | 2.66 (2.15 – 3.17) | 3.53 (2.36 - 5.26) | 5.20 (3.48 – 6.91) |
| 2514 | 1.66 (1.16 - 2.37) | 2.19 (1.51 – 2.87) | 1.54 (0.64 - 3.69) | 1.86 (0.38 – 3.34) |
| 2515 | 1.79 (1.48 - 2.17) | 3.14 (2.69 – 3.59) | 7.87 (6.35 - 9.76) | 13.60 (11.41 – 15.87) |
| 2516 | 1.52 (1.21 - 1.91) | 3.22 (2.72 – 3.73) | 3.52 (2.50 - 4.95) | 7.15 (5.44 – 8.87) |
| 2517 | 2.45 (2.05 - 2.94) | 2.75 (2.28 – 3.21) | 6.55 (5.06 - 8.47) | 8.12 (6.25 – 10.00) |
| 2518 | 1.24 (0.98 - 1.57) | 2.31 (1.91 – 2.72) | 4.53 (3.36 - 6.11) | 9.41 (7.46 – 11.36) |
| 2521 | 1.66 (1.33 - 2.08) | 3.32 (2.79 – 3.84) | 2.79 (1.90 - 4.10) | 5.87 (4.31 – 7.43) |
| 2522 | 1.53 (1.24 - 1.87) | 2.92 (2.49 – 3.35) | 3.26 (2.33 - 4.56) | 5.95 (4.47 – 7.42) |
| 2523 | 1.50 (1.09 - 2.05) | 3.50 (2.78 – 4.21) | 2.87 (1.67 - 4.94) | 4.30 (2.39 – 6.21) |
| 2524 | 1.72 (1.32 - 2.25) | 3.73 (3.05 – 4.40) | 4.80 (3.24 - 7.10) | 8.09 (5.65 – 10.53) |
| 2525 | 2.15 (1.83 - 2.53) | 3.77 (3.31 – 4.22) | 9.44 (7.84 - 1.36) | 13.40 (11.29 – 15.45) |
| 2526 | 2.70 (2.31 - 3.15) | 4.22 (3.69 – 4.74) | 9.09 (7.40 - 1.16) | 12.30 (10.10 – 15.45) |
| 2531 | 2.45 (2.02 - 2.98) | 3.47 (2.90 – 4.03) | 6.03 (4.46 - 8.16) | 8.44 (6.28 – 10.60) |
| 2532 | 2.77 (2.20 - 3.50) | 3.45 (2.73 – 4.17) | 3.80 (2.37 - 6.12) | 6.52 (4.15 – 8.89) |
| 2533 | 2.02 (1.59 - 2.57) | 2.32 (1.81 – 2.84) | 5.01 (3.48 - 7.21) | 5.65 (3.72 – 7.59) |
| 2541 | 1.57 (1.21 - 2.04) | 2.40 (1.90 – 2.90) | 5.03 (3.55 - 7.11) | 7.35 (5.24 – 9.45) |
| 2542 | 3.73 (3.21 - 4.33) | 3.78 (3.22 – 4.34) | 7.97 (6.22 - 10.20) | 8.80 (6.73 – 10.87) |
| 2543 | 2.13 (1.67 - 2.73) | 2.75 (2.15 – 3.35) | 4.01 (2.59 - 6.21) | 4.76 (2.84 – 6.67) |
| 2544 | 2.37 (1.99 - 2.82) | 2.33 (1.92 – 2.74) | 7.82 (6.23 - 9.82) | 8.44 (6.59 – 10.30) |
| 2545 | 1.87 (1.54 - 2.28) | 2.10 (1.72 – 2.49) | 4.63 (3.44 - 6.22) | 5.44 (3.96 – 6.92) |
| 2546 | 1.52 (1.13 - 2.05) | 2.09 (1.57 – 2.62) | 2.99 (1.80 - 4.96) | 4.23 (2.43 – 6.03) |
| 2547 | 3.38 (2.84 – 4.03) | 2.64 (2.12 – 3.16) | 5.23 (3.74 - 7.32) | 4.97 (3.25 – 6.68) |
| 2548 | 1.57 (1.33 - 1.84) | 2.19 (1.89 – 2.49) | 8.05 (6.80 - 9.53) | 11.80 (10.11 – 13.39) |
| 2551 | 3.58 (3.12 - 4.12) | 2.59 (2.17 – 3.02) | 6.78 (5.29 - 8.70) | 5.03 (3.58 – 6.49) |
| 2552 | 3.35 (2.97 - 3.79) | 2.87 (2.50 – 3.25) | 7.59 (6.25 - 9.22) | 5.98 (4.67 – 7.28) |
| 2553 | 2.90 (2.42 - 3.48) | 2.50 (2.02 – 2.99) | 5.01 (3.66 - 6.86) | 4.60 (3.10 – 6.04) |
| 2554 | 2.30 (1.52 - 3.50) | 1.55 (0.76 – 2.34) | 8.82 (5.32 - 14.64) | 6.61 (2.74 – 10.47) |
| 2555 | 3.01 (2.55 - 3.55) | 1.68 (1.32 – 2.05) | 7.93 (6.21 - 10.13) | 4.57 (3.09 – 6.04) |
| 2561 | 1.01 (0.69 - 1.47) | 2.02 (1.48 – 2.56) | 3.58 (2.19 - 5.84) | 4.84 (2.80 – 6.87) |
| 2562 | 1.59 (1.29 - 1.97) | 2.84 (2.39 – 3.28) | 5.99 (4.61 - 7.78) | 9.94 (7.92 – 11.97) |
| 2563 | 1.18 (0.92 - 1.53) | 1.96 (1.57 – 2.35) | 1.99 (1.24 - 3.21) | 3.28 (2.07 – 4.50) |
| 2564 | 1.56 (1.26 - 1.93) | 1.68 (1.33 – 2.03) | 4.40 (3.23 - 6.00) | 5.05 (3.59 – 6.51) |
| 2565 | 1.61 (1.32 - 1.97) | 1.51 (1.19 – 1.82) | 3.45 (2.46 - 4.82) | 3.33 (2.19 – 4.47) |
| 2566 | 2.31 (1.82 - 2.93) | 1.65 (1.19 – 2.12) | 6.74 (4.81 - 9.43) | 4.06 (2.30 – 5.82) |
| 2571 | 2.21 (1.83 - 2.66) | 3.22 (2.72 – 3.73) | 6.50 (5.00 - 8.45) | 9.60 (7.53 – 11.67) |
| 2572 | 2.28 (1.87 - 2.78) | 3.89 (3.30 – 4.48) | 11.47 (9.27 - 14.18) | 18.00 (14.91 – 21.01) |
| 2573 | 1.21 (0.93 - 1.56) | 2.46 (2.02 – 2.90) | 2.03 (1.26 - 3.26) | 3.97 (2.62 -5.32 ) |
| 2574 | 1.77 (1.37 - 2.27) | 2.63 (2.09 – 3.17) | 2.71 (1.66 - 4.42) | 4.45 (2.75 – 6.15) |
| 2581 | 1.69 (1.14 - 2.49) | 2.05 (1.33 – 2.78) | 2.81 (1.34 - 5.90) | 4.70 (2.01 – 7.39) |
| 2582 | 1.44 (1.12 - 1.87) | 1.66 (1.26 – 2.05) | 1.59 (0.88 - 2.87) | 1.79 (0.79 – 2.79) |
| 2583 | 2.71 (2.30 - 3.19) | 2.50 (2.08 – 2.93) | 4.85 (3.62 - 6.49) | 4.86 (3.44 – 6.28) |
| 2584 | 3.40 (2.83 - 4.08) | 2.98 (2.39 – 3.56) | 6.93 (5.09 - 9.45) | 5.61 (3.68 – 7.55) |
| 2585 | 2.16 (1.71 - 2.73) | 1.90 (1.43 – 2.38) | 2.88 (1.77 - 4.70) | 2.71 (1.34 – 4.08) |
| 2586 | 2.43 (1.99 - 2.97) | 2.45 (1.96 – 2.94) | 3.74 (2.53 - 5.53) | 3.65 (2.20 – 5.10) |
| 2587 | 2.25 (1.82 - 2.77) | 2.13 (1.67 – 2.59) | 5.64 (4.11 - 7.76) | 5.29 (3.55 – 7.03) |
| 2591 | 3.24 (2.69 - 3.90) | 2.86 (2.29 – 3.43) | 4.51 (3.09 - 6.57) | 4.73 (2.99 – 6.47) |
| 2592 | 3.04 (2.50 - 3.69) | 2.27 (1.75 – 2.78) | 6.60 (4.80 - 9.07) | 4.99 (3.17 – 6.81) |
| 2593 | 1.61 (1.28 - 2.03) | 2.39 (1.94 – 2.85) | 2.83 (1.84 - 4.33) | 4.29 (2.80 – 5.78) |
| 2594 | 3.01 (2.09 - 4.33) | 1.75 (0.91 – 2.58) | 11.86 (7.65 - 18.39) | 7.57 (3.41 – 11.72) |
| 2595 | 1.03 (0.75 - 1.40) | 1.72 (1.31 – 2.13) | 2.61 (1.66 - 4.09) | 4.51 (2.96 – 6.05) |
| 2596 | 2.04 (1.61 - 2.59) | 1.66 (1.22 – 2.10) | 6.21 (4.50 - 8.57) | 5.12 (3.30 – 6.94) |
| 2597 | 3.45 (2.92 - 4.07) | 1.55 (1.16 – 1.93) | 8.65 (6.73 - 11.12) | 3.51 (2.13 – 4.90) |

Postal codes 2491 and 2495 were omitted due to potential personal data disclosure.

**Table S2 – Estimated average household SES percentile by neighbourhood in The Hague in 2014-2019 and 2020.**

| **Postal code** | **Cohort 2014 – 2019**  **(SD)*** | **Cohort 2020**  **(SD)*** |
| --- | --- | --- |
| 2492 | 61.53 (25.64) | 63.59 (25.99) |
| 2493 | 60.38 (25.28) | 63.75 (25.49) |
| 2496 | 60.31 (28.51) | 60.95 (28.95) |
| 2497 | 55.03 (27.16) | 56.60 (27.01) |
| 2498 | 63.36 (28.00) | 66.03 (28.20) |
| 2511 | 45.93 (31.70) | 46.70 (31.81) |
| 2512 | 33.94 (27.79) | 35.23 (27.91) |
| 2513 | 45.99 (31.43) | 45.68 (31.78) |
| 2514 | 61.88 (33.85) | 60.90 (33.77) |
| 2515 | 31.62 (27.06) | 32.12 (27.32) |
| 2516 | 23.68 (22.37) | 27.50 (25.07) |
| 2517 | 61.66 (32.89) | 62.03 (33.00) |
| 2518 | 48.97 (32.54) | 51.02 (32.36) |
| 2521 | 29.86 (25.69) | 29.81 (25.80) |
| 2522 | 35.68 (23.58) | 36.27 (23.32) |
| 2523 | 36.00 (23.33) | 37.73 (23.44) |
| 2524 | 35.54 (24.35) | 37.34 (24.87) |
| 2525 | 30.68 (23.50) | 34.21 (24.93) |
| 2526 | 26.14 (21.70) | 27.86 (22.77) |
| 2531 | 31.96 (23.60) | 33.67 (24.58) |
| 2532 | 28.25 (22.18) | 28.73 (22.96) |
| 2533 | 27.36 (21.73) | 27.92 (22.37) |
| 2541 | 30.69 (22.75) | 31.58 (23.31) |
| 2542 | 32.58 (22.42) | 33.21 (22.77) |
| 2543 | 33.29 (24.30) | 33.46 (24.67) |
| 2544 | 33.93 (23.77) | 35.60 (25.48) |
| 2545 | 42.29 (26.55) | 42.87 (26.80) |
| 2546 | 48.07 (23.22) | 48.21 (23.58) |
| 2547 | 44.83 (23.07) | 44.26 (23.01) |
| 2548 | 58.06 (27.17) | 59.84 (27.58) |
| 2551 | 46.30 (25.48) | 45.36 (25.59) |
| 2552 | 52.19 (26.68) | 51.76 (26.72) |
| 2553 | 53.82 (28.80) | 56.42 (29.26) |
| 2554 | 71.39 (28.18) | 70.89 (28.77) |
| 2555 | 56.34 (27.47) | 55.92 (27.80) |
| 2561 | 53.58 (29.34) | 55.45 (29.55) |
| 2562 | 41.03 (28.81) | 42.99 (28.96) |
| 2563 | 48.29 (27.85) | 49.95 (28.27) |
| 2564 | 62.13 (26.12) | 63.92 (25.74) |
| 2565 | 60.48 (26.51) | 61.67 (26.24) |
| 2566 | 79.60 (23.84) | 80.92 (23.03) |
| 2571 | 31.64 (23.84) | 33.75 (24.44) |
| 2572 | 31.96 (23.27) | 35.78 (24.28) |
| 2573 | 37.57 (23.39) | 39.25 (23.61) |
| 2574 | 41.33 (23.81) | 41.99 (23.78) |
| 2581 | 65.12 (25.68) | 65.56 (26.23) |
| 2582 | 73.89 (28.58) | 74.37 (28.84) |
| 2583 | 44.84 (27.13) | 46.80 (27.98) |
| 2584 | 51.80 (30.24) | 51.89 (31.23) |
| 2585 | 68.54 (31.61) | 69.44 (31.91) |
| 2586 | 55.70 (30.02) | 56.44 (30.12) |
| 2587 | 62.79 (32.66) | 64.80 (32.11) |
| 2591 | 43.40 (28.37) | 43.22 (28.45) |
| 2592 | 42.01 (27.34) | 42.29 (27.33) |
| 2593 | 55.14 (28.84) | 56.49 (28.79) |
| 2594 | 73.16 (28.79) | 71.55 (30.27) |
| 2595 | 53.89 (32.48) | 53.33 (32.81) |
| 2596 | 78.42 (26.52) | 79.07 (26.91) |
| 2597 | 78.15 (25.87) | 77.95 (26.17) |

*Standard Deviation. Postal codes 2491 and 2495 were omitted due to potential personal data disclosure.

**Table S3 – Univariable, multivariable with structural determinants, and full multivariable Poisson regression analyses of determinants associated with complicated LRTI incidence, between the ages of 18 and 80, in 2014-2019.**

|  | **Jan 2014 – Dec 2019** | | |
| --- | --- | --- | --- |
| **Independent Variables** | **IRR^a^ (95% CI)** | **aIRR^b^ (95% CI)**  **[structural determinants]** | **aIRR^c^ (95% CI)**  **[all determinants]** |
| **Sex** |  | | |
| Male | **Ref** | **Ref** | **Ref** |
| Female | 0.92 (0.85 – 0.98) | 0.85 (0.79 – 0.92) | 0.84 (0.78 – 0.90) |
| **Age^d^** |  | | |
| 18-49 | **Ref** | **Ref** | **Ref** |
| 50 – 59 | 2.99 (2.65 – 3.37) | 2.97 (2.63 – 3.37) | 2.20 (1.93 – 2.49) |
| 60 – 69 | 7.27 (6.54 – 8.07) | 7.18 (6.44 – 8.00) | 4.28 (3.79 – 4.83) |
| 70 – 80 | 15.82 (14.33 – 17.46) | 14.76 (13.28 – 16.41) | 7.28 (6.43 – 8.25) |
| **SES (quintiles where 5 is the lowest)** |  | | |
| 1 | **Ref** | **Ref** | **Ref** |
| 2 | 1.07 (0.92 – 1.24) | 1.35 (1.16 – 1.56) | 1.18 (1.02 – 1.37) |
| 3 | 1.41 (1.23 – 1.62) | 1.78 (1.55 – 2.05) | 1.47 (1.28 – 1.69) |
| 4 | 1.99 (1.75 – 2.27) | 2.15 (1.89 – 2.45) | 1.59 (1.39 – 1.81) |
| 5 | 2.48 (2.20 – 2.80) | 3.47 (3.06 – 3.94) | 2.26 (1.98 – 2.57) |
| **Migration Background** |  | | |
| The Netherlands | **Ref** | **Ref** | **Ref** |
| Middle & Eastern Europe | 0.20 (0.15 – 0.28) | 0.42 (0.30 – 0.58) | 0.51 (0.37 – 0.72) |
| Other Europe | 0.56 (0.48 – 0.66) | 0.75 (0.64 – 0.87) | 0.81 (0.69 – 0.94) |
| Turkey | 0.69 (0.59 – 0.81) | 1.00 (0.85 – 1.17) | 0.99 (0.84 – 1.16) |
| Morocco | 0.88 (0.74 – 1.04) | 1.00 (0.84 – 1.18) | 1.10 (0.93 – 1.30) |
| Suriname | 1.12 (1.00 – 1.24) | 1.23 (1.10 – 1.38) | 1.08 (0.96 – 1.21) |
| Dutch Caribbean | 0.49 (0.37 – 0.66) | 0.64 (0.47 – 0.86) | 0.72 (0.53 – 0.97) |
| Indonesia | 0.76 (0.63 – 0.92) | 0.72 (0.60 – 0.87) | 0.78 (0.65 – 0.94) |
| Other Africa | 0.39 (0.30 – 0.51) | 0.57 (0.43 – 0.76) | 0.62 (0.47 – 0.83) |
| Other Asia | 0.42 (0.33 – 0.52) | 0.59 (0.47 – 0.74) | 0.68 (0.54 – 0.86) |
| Other America’s and Oceania | 0.34 (0.24 – 0.49) | 0.61 (0.42 – 0.88) | 0.70 (0.49 – 1.01) |
| **Household Composition** |  | | |
| One person household without children | **Ref** | **-** | **Ref** |
| One parent household with children | 0.46 (0.39 – 0.54) | - | 0.90 (0.77 – 1.07) |
| Couple without children | 1.00 (0.92 – 1.10) | - | 0.93 (0.85 – 1.02) |
| Couple with children | 0.36 (0.32 – 0.40) | - | 0.97 (0.86 – 1.09) |
| More than two-person adult household without children | 0.49 (0.34 – 0.71) | - | 0.84 (0.58 – 1.23) |
| More than two-person adult household with children | 0.86 (0.68 – 1.07) | - | 1.40 (1.12 – 1.75) |
| Institutional household | 6.46 (5.66 – 7.37) | - | 4.11 (3.49 – 4.82) |
| Other household | 0.50 (0.30 – 0.81) | - | 1.16 (0.72 – 1.86) |
| **Comorbidities** |  | | |
| Neoplastic Disease | 1.95 (1.79– 2.13) | **-** | 1.54 (1.41 – 1.68) |
| Liver Disease | 3.38 (2.69 – 4.26) | - | 1.76 (1.40 – 2.21) |
| Congestive Heart Failure | 13.14 (11.87 – 14.55) | - | 2.25 (1.99 – 2.55) |
| Cerebrovascular Disease | 4.94 (4.34 – 5.63) | - | 1.30 (1.13 – 1.49) |
| Renal Disease | 8.76 (7.68 – 9.99) | - | 1.73 (1.50 – 2.00) |
| Pulmonary Disease | 13.17 (12.22 – 14.21) | - | 5.60 (5.13 – 6.11) |
| Dementia | 5.66 (4.54 – 7.05) | - | 1.09 (0.87 – 1.36) |
| Neurologic Disease | 4.64 (3.71 – 5.80) | -- | 2.25 (1.79 – 2.84) |
| Diabetes mellitus | 3.89 (3.57 – 4.23) | - | 1.46 (1.33 – 1.60) |
| HIV | 2.88 (1.96 – 4.24) | - | 3.50 (2.36 – 5.20) |
| Immunocompromised | 3.84 (3.47 – 4.26) | - | 2.25 (2.02 – 2.51) |
| Other Cardiovascular Disease | 5.01 (4.58 – 5.48) | - | 1.08 (0.97 – 1.20) |
| **Flu season** |  | | |
| 13/14 | **Ref** | **Ref** | **Ref** |
| 14/15 | 1.12 (0.96 – 1.30) | 1.44 (1.23 – 1.69) | 1.43 (1.22 – 1.68) |
| 15/16 | 0.92 (0.79 – 1.07) | 1.18 (1.01 – 1.39) | 1.18 (1.00 – 1.38) |
| 16/17 | 1.05 (0.91 – 1.22) | 1.33 (1.14 – 1.55) | 1.34 (1.15 – 1.57) |
| 17/18 | 1.23 (1.06 – 1.42) | 1.55 (1.33 – 1.81) | 1.61 (1.37 – 1.87) |
| 18/19 | 1.19 (1.03 – 1.38) | 1.48 (1.26 – 1.72) | 1.58 (1.36 – 1.85) |
| 19/20 | 1.07 (0.90 – 1.26) | 1.86 (1.54 – 2.25) | 2.06 (1.70 – 2.50) |
| **Quartile** |  | | |
| 1 | **Ref** | **Ref** | **Ref** |
| 2 | 0.55 (0.50– 0.60) | 0.55 (0.50 – 0.60) | 0.53 (0.48 – 0.58) |
| 3 | 0.43 (0.39 – 0.47) | 0.38 (0.35 – 0.43) | 0.37 (0.33 – 0.41) |
| 4 | 0.56 (0.51 – 0.61) | 0.50 (0.46 – 0.56) | 0.48 (0.44 – 0.53) |
| **Complicated LRTI episode in the past year** |  | | |
| No | **Ref** | **Ref** | **Ref** |
| Yes | 24.59 (19.95 – 30.33) | 7.55 (6.01 – 9.49) | 2.62 2.05 – 3.34) |

Flu season was defined based on yearly quartiles where a flu season ranged from 1 July until 30 June the following year. Flu season 13/14 encompassed 1 January 2014 until 30 June 2014, flu season 19/20 encompassed 1 July 2019 until 31 December 2019. **a**. Univariable unadjusted incidence rate ratio **b.** Adjusted Incidence rate ratio for a multivariable model with structural determinants SES, Migration background, age, and sex, and flu season, quartile of a year, and LRTI hospitalisation in the past year **c.** Adjusted Incidence rate ratio for the full multivariable model with SES, migration background, age, sex, household composition, comorbidity, flu season, quartile of a year, and LRTI hospitalisation in the past year. **d.** Those above 80 years of age were excluded from our regression analyses due to the small number of individuals over 80 years of age in the migration background groups

**Table S4 – Univariable, multivariable with structural determinants, and full multivariable Poisson regression analyses of determinants associated with complicated LRTI incidence, between the ages of 18 and 80, in 2020.**

|  | **Jan 2020 – Dec 2020** | | |
| --- | --- | --- | --- |
| **Independent Variables** | **IRR^a^ (95% CI)** | **aIRR^b^ (95% CI)**  **[structural determinants]** | **aIRR^c^ (95% CI)**  **[all determinants]** |
| **Sex** |  | | |
| Male | **Ref** | **Ref** | **Ref** |
| Female | 0.66 (0.59 – 0.73) | 0.64 (0.57 – 0.70) | 0.64 (0.57 - 0.71) |
| **Age^d^** |  | | |
| 18-49 | **Ref** | **Ref** | **Ref** |
| 50 – 59 | 4.08 (3.47 – 4.79) | 4.36 (3.70 – 5.13) | 3.36 (2.84 – 3.98) |
| 60 – 69 | 7.50 (6.45 – 8.73) | 8.42 (7.21 – 9.84) | 5.45 (4.58 – 6.48) |
| 70 – 80 | 12.96 (11.19 – 15.00) | 15.61 (13.38 – 18.21) | 8.20 (6.82 – 9.86) |
| **SES (quintiles where 5 is the lowest)** |  | | |
| 1 | **Ref** | **Ref** | **Ref** |
| 2 | 1.24 (1.02 – 1.52) | 1.38 (1.13 – 1.68) | 1.26 (1.03 – 1.54) |
| 3 | 1.50 (1.24 – 1.81) | 1.60 (1.32 – 1.93) | 1.39 (1.15 – 1.69) |
| 4 | 1.78 (1.49 – 2.13) | 1.69 (1.41 – 2.04) | 1.38 (1.14 – 1.66) |
| 5 | 2.36 (2.00 – 2.78) | 1.99 (1.66 – 2.38) | 1.47 (1.21 – 1.77) |
| **Migration Background** |  |  |  |
| The Netherlands | **Ref** | **Ref** | **Ref** |
| Middle & Eastern Europe | 0.31 (0.21 – 0.46) | 0.83 (0.56 – 1.25) | 0.99 (0.66 – 1.48) |
| Other Europe | 0.65 (0.51 – 0.84) | 1.04 (0.81 – 1.33) | 1.12 (0.87 – 1.44) |
| Turkey | 1.96 (1.65 – 2.31) | 3.36 (2.81 – 4.02) | 2.83 (2.35 – 3.40) |
| Morocco | 2.46 (2.07 – 2.93) | 3.53 (2.93 – 4.25) | 3.19 (2.63 – 3.89) |
| Suriname | 2.19 (1.89 – 2.54) | 2.60 (2.23 – 3.02) | 2.00 (1.71 – 2.34) |
| Dutch Caribbean | 1.04 (0.74 – 1.45) | 1.68 (1.19 – 2.37) | 1.69 (1.19 – 2.40) |
| Indonesia | 1.57 (1.23 – 2.01) | 1.35 (1.05 – 1.73) | 1.42 (1.11 – 1.83) |
| Other Africa | 1.27 (0.97 – 1.67) | 2.15 (1.63 – 2.83) | 2.07 (1.57 – 2.74) |
| Other Asia | 0.77 (0.60 – 0.99) | 1.31 (1.01 – 1.71) | 1.34 (1.03 – 1.75) |
| Other America’s and Oceania | 0.57 (0.37 – 0.88) | 1.22 (0.79 – 1.88) | 1.35 (0.87 – 2.09) |
| **Household Composition** |  | | |
| One person household without children | **Ref** | **-** | **Ref** |
| One parent household with children | 0.64 (0.51 – 0.82) | - | 1.04 (0.82 – 1.33) |
| Couple without children | 1.36 (1.19 – 1.56) | - | 1.15 (1.00 – 1.32) |
| Couple with children | 0.80 (0.69 – 0.92) | - | 1.41 (1.20 – 1.66) |
| More than two-person adult household without children | 0.72 (0.44 – 1.18) | - | 0.88 (0.54 – 1.44) |
| More than two-person adult household with children | 1.44 (1.07 – 1.94) | - | 1.47 (1.08 – 2.01) |
| Institutional household | 8.19 (6.79 – 9.89) | - | 6.69 (5.42 – 8.27) |
| Other household | 0.63 (0.28 – 1.41) | - | 1.11 (0.50 – 2.49) |
| **Comorbidities** |  | | |
| Neoplastic Disease | 1.58 (1.40 – 1.80) | **-** | 1.51 (1.32 – 1.72) |
| Liver Disease | 3.42 (2.54 – 4.62) | - | 1.71 (1.27 – 2.30) |
| Congestive Heart Failure | 10.90 (9.16 – 12.97) | - | 2.32 (1.89 – 2.85) |
| Cerebrovascular Disease | 5.35 (4.43 – 6.46) | - | 1.50 (1.23 – 1.83) |
| Renal Disease | 7.79 (6.28 – 9.65) | -- | 1.41 (1.12 – 1.78) |
| Pulmonary Disease | 5.94 (5.21 – 6.77) | - | 2.58 (2.24 – 2.98) |
| Dementia | 7.76 (5.74 – 10.49) | - | 1.31 (0.95 – 1.80) |
| Neurologic Disease | 4.30 (3.10 – 5.95) | -- | 2.20 (1.55 – 3.13) |
| Diabetes mellitus | 6.42 (5.77 – 7.16) | - | 2.06 (1.82 – 2.34) |
| HIV | 1.12 (0.47 – 2.69) | - | 1.21 (0.50 – 2.92) |
| Immunocompromised | 6.42 (5.65 – 7.28) | - | 3.96 (3.46 – 4.53) |
| Other Cardiovascular Disease | 5.02 (4.42 – 5.71) | - | 1.16 (1.00 – 1.35) |
| **Quartile** |  | | |
| 1 | **Ref** | **Ref** | **Ref** |
| 2 | 1.15 (0.99 – 1.34) | 1.13 (0.97 – 1.32) | 1.14 (0.98 – 1.33) |
| 3 | 0.79 (0.67 – 0.93) | 0.77 (0.65 – 0.91) | 0.78 (0.66 – 0.93) |
| 4 | 2.05 (1.79 – 2.35) | 2.00 (1.75 – 2.30) | 2.07 (1.80 – 2.37) |
| **Complicated LRTI episode in the past year** |  | | |
| No | **Ref** | **Ref** | **Ref** |
| Yes | 10.49 (7.24 – 15.21) | 3.59 (2.46 – 5.24) | 1.51 (1.04 – 2.30) |

**a**. Univariable unadjusted incidence rate ratio **b.** Adjusted Incidence rate ratio for a multivariable model with structural determinants SES, Migration background, age, and sex, and quartile of a year, and LRTI hospitalisation in the past year **c.** Adjusted Incidence rate ratio for the full multivariable model with SES, migration background, age, sex, household composition, comorbidity, quartile of a year, and LRTI hospitalisation in the past year. **d.** Those above 80 years of age were excluded from our regression analyses due to the small number of individuals over 80 years of age in the migration background groups

**Table S5 – Univariable, multivariable with structural determinants, and full multivariable Poisson regression analyses of determinants associated with LRTI-associated mortality, between the ages of 18 and 80, in 2014-2019.**

|  | **Jan 2014 – Dec 2019** | | |
| --- | --- | --- | --- |
| **Independent Variables** | **IRR^a^ (95% CI)** | **aIRR^b^ (95% CI)**  **[structural determinants]** | **aIRR^c^ (95% CI)**  **[all determinants]** |
| **Sex** |  | | |
| Male | **Ref** | **Ref** | **Ref** |
| Female | 0.73 (0.60 – 0.90) | 0.62 (0.50 – 0.76) | 0.64 (0.51 – 0.80) |
| **Age^e^** |  | | |
| 18-49 | 0.01 (0.01 – 0.02) | 0.01 (0.01 – 0.02) | 0.02 (0.01 – 0.03) |
| 50 – 59 | 0.08 (0.06 – 0.12) | 0.10 (0.07 – 0.14) | 0.15 (0.10 – 0.23) |
| 60 – 69 | 0.32 (0.25 – 0.44) | 0.35 (0.27 – 0.44) | 0.43 (0.33 – 0.55) |
| 70 – 80 | **Ref** | **Ref** | **Ref** |
| **SES (quintiles where 5 is the lowest)** |  | | |
| 1 | **Ref** | **Ref** | **Ref** |
| 2 | 1.02 (0.62 – 1.67) | 1.48 (0.90 – 2.42) | 1.29 (0.79 – 2.13) |
| 3 | 1.81 (1.18 – 2.78) | 2.63 (1.70 – 4.06) | 2.16 (1.39 – 3.37) |
| 4 | 2.88 (1.94 – 4.28) | 3.31 (2.20 – 4.97) | 2.33 (1.53 – 3.56) |
| 5 | 3.22 (2.21 – 4.70) | 6.17 (4.12 – 9.24) | 3.49 (2.28 – 5.34) |
| **Migration Background** |  | | |
| The Netherlands | **Ref** | **Ref** | **Ref** |
| Middle & Eastern Europe | 0.14 (0.05 – 0.38) | 0.48 (0.15 – 1.51) | 0.60 (0.19 – 1.91) |
| Other Europe | 0.62 (0.42 – 0.91) | 0.85 (0.56 – 1.29) | 0.92 (0.61 - 1.40) |
| Turkey | 0.20 (0.10 – 0.41) | 0.35 (0.17 – 0.70) | 0.42 (0.20 – 0.86) |
| Morocco | 0.35 (0.18 – 0.65) | 0.41 (0.21 – 0.77) | 0.57 (0.30 – 1.11) |
| Suriname | 0.61 (0.43 – 0.88) | 0.71 (0.48 – 1.04) | 0.73 (0.49 – 1.08) |
| Dutch Caribbean | 0.21 (0.07 – 0.67) | 0.24 (0.06 – 0.95) | 0.26 (0.06 – 1.06) |
| Indonesia | 0.77 (0.47 – 1.26) | 0.69 (0.41 – 1.16) | 0.74 (0.44 – 1.25) |
| Other Africa^d^ | - | - | - |
| Other Asia | 0.35 (0.19 – 0.63) | 0.55 (0.28 – 1.07) | 0.74 (0.38 – 1.44) |
| Other America’s and Oceania | 0.42 (0.19 – 0.95) | 0.92 (0.38 – 2.24) | 1.15 (0.47 – 2.82) |
| **Household Composition** |  | | |
| One person household without children | **Ref** | **-** | **Ref** |
| One parent household with children | 0.17 (0.09 – 0.36) | - | 0.65 (0.31 – 1.33) |
| Couple without children | 0.99 (0.78 – 1.25) | - | 0.86 (0.66 – 1.10) |
| Couple with children | 0.13 (0.09 – 0.21) | - | 0.71 (0.44 – 1.15) |
| More than two-person adult household without children | 0.26 (0.07 – 1.07) | - | 0.65 (0.16 – 2.61) |
| More than two-person adult household with children | 0.49 (0.22 – 1.12) | - | 1.36 (0.60 – 3.10) |
| Institutional household | 12.70 (9.60 – 16.81) | - | 7.74 (5.51 – 10.86) |
| Other household | 0.26 (0.04 – 1.86) | - | 0.98 (0.14 – 7.02) |
| **Comorbidities** |  | | |
| Neoplastic Disease | 2.25 (1.79 – 2.82) | **-** | 1.66 (1.30 – 2.11) |
| Liver Disease | 3.59 (2.02 – 6.38) | - | 2.00 (1.13 – 3.54) |
| Congestive Heart Failure | 20.50 (16.02 – 26.24) | - | 3.14 (2.30 – 4.28) |
| Cerebrovascular Disease | 4.74 (3.29 – 6.85) | - | 0.89 (0.60 – 1.32) |
| Renal Disease | 9.35 (6.55 – 13.36) | - | 1.50 (1.01 – 2.22) |
| Pulmonary Disease | 13.69 (11.11 – 16.87) | -- | 4.77 (3.75 – 6.06) |
| Dementia | 7.84 (4.60 – 13.35) | - | 1.06 (0.61 – 1.82) |
| Neurologic Disease | 7.52 (4.63 – 12.22) | - | 2.63 (1.56 – 4.41) |
| Diabetes mellitus | 3.65 (2.88 – 4.63) | - | 1.16 (0.90 – 1.50) |
| HIV | 1.77 (0.44 – 7.08) | - | 2.36 (0.58 – 9.57) |
| Immunocompromised | 1.83 (1.23 – 2.73) | - | 1.09 (0.72 – 1.64) |
| Other Cardiovascular Disease | 5.44 (4.28 – 6.92) | - | 0.89 (0.67 – 1.18) |
| **Flu season** |  | | |
| 13/14 | **Ref** | **Ref** | **Ref** |
| 14/15 | 1.02 (0.64 – 1.61) | 1.42 (0.87 – 2.32) | 1.43 (0.88 – 2.34) |
| 15/16 | 0.86 (0.54 – 1.38) | 1.11 (0.67 – 1.84) | 1.12 (0.67 – 1.87) |
| 16/17 | 0.80 (0.50 – 1.29) | 1.11 (0.67 – 1.82) | 1.14 (0.69 – 1.88) |
| 17/18 | 1.05 (0.67 – 1.66) | 1.38 (0.85 – 2.24) | 1.45 (0.89 – 2.36) |
| 18/19 | 1.78 (1.17 – 2.72) | 2.36 (1.49 – 3.73) | 2.54 (1.59 – 4.04) |
| 19/20 | 1.60 (1.00 – 2.57) | 3.73 (2.15 – 6.47) | 4.10 (2.34 – 7.17) |
| **Quartile** |  | | |
| 1 | **Ref** | **Ref** | **Ref** |
| 2 | 0.53 (0.41 – 0.69) | 0.54 (0.41 – 0.71) | 0.54 (0.41 – 0.71) |
| 3 | 0.44 (0.33 – 0.58) | 0.33 (0.24 – 0.45) | 0.32 (0.24 – 0.45) |
| 4 | 0.42 (0.32 – 0.56) | 0.33 (0.24 – 0.45) | 0.32 (0.23 – 0.44) |
| **Complicated LRTI episode in the past year** |  | | |
| No | **Ref** | **Ref** | **Ref** |
| Yes | 41.25 (25.38 – 67.05) | 9.16 (5.39 – 15.58) | 3.04 (1.76 – 5.26) |

Flu season was defined based on yearly quartiles where a flu season ranged from 1 July until 30 June the following year. Flu season 13/14 encompassed 1 January 2014 until 30 June 2014, flu season 19/20 encompassed 1 July 2019 until 31 December 2019. **a**. Univariable unadjusted incidence rate ratio **b.** Adjusted Incidence rate ratio for a multivariable model with structural determinants SES, Migration background, age, and sex, and flu season, quartile of a year, and LRTI hospitalisation in the past year **c.** Adjusted Incidence rate ratio for the full multivariable model with SES, migration background, age, sex, household composition, comorbidity, flu season, quartile of a year, and LRTI hospitalisation in the past year. **d.** ‘Other Africa’ was not included in analysis due to insufficient cases. **e.** Those above 80 years of age were excluded from our regression analyses due to the small number of individuals over 80 years of age in the migration background groups

**Table S6 – Univariable, multivariable with structural determinants, and full multivariable Poisson regression analyses of determinants associated with LRTI-associated mortality, between the ages of 18 and 80, in 2020.**

|  | **Jan 2020 – Dec 2020** | | |
| --- | --- | --- | --- |
| **Independent Variables** | **IRR^a^ (95% CI)** | **aIRR^b^ (95% CI)**  **[structural determinants]** | **aIRR^c^ (95% CI)**  **[all determinants]** |
| **Sex** |  | | |
| Male | **Ref** | **Ref** | **Ref** |
| Female | 0.60 (0.48 – 0.75) | 0.53 (0.42 – 0.67) | 0.61 (0.48 - 0.77) |
| **Age^e^** |  | | |
| 18-49 | 0.01 (0.01 – 0.02) | 0.01 (0.01 – 0.02) | 0.02 (0.01 – 0.03) |
| 50 – 59 | 0.06 (2.78 – 9.87) | 0.06 (0.04 – 0.09) | 0.09 (0.06 – 0.15) |
| 60 – 69 | 0.26 (0.20 – 0.34) | 0.27 (0.21 – 0.36) | 0.36 (0.27 – 0.48) |
| 70 – 80 | **Ref** | **Ref** | **Ref** |
| **SES (quintiles where 5 is the lowest)** |  | | |
| 1 | **Ref** | **Ref** | **Ref** |
| 2 | 1.51 (0.89 – 2.54) | 1.99 (1.18 – 3.37) | 1.79 (1.05 – 3.03) |
| 3 | 1.87 (1.13 – 3.07) | 2.39 (1.45 – 3.94) | 1.96 (1.18 – 3.25) |
| 4 | 3.09 (1.96 – 4.88) | 3.29 (2.06 – 5.25) | 2.17 (1.34 – 3.53) |
| 5 | 4.33 (2.82 – 6.64) | 4.98 (3.18 – 7.80) | 2.59 (1.61 – 4.16) |
| **Migration Background** |  | | |
| The Netherlands | **Ref** | **Ref** | **Ref** |
| Middle & Eastern Europe | 0.14 (0.04 – 0.43) | 0.48 (0.12 – 1.94) | 0.59 (0.14 – 2.43) |
| Other Europe | 0.71 (0.45 – 1.13) | 1.31 (0.82 – 2.10) | 1.40 (0.87 – 2.26) |
| Turkey | 0.72 (0.44 – 1.17) | 1.43 (0.86 – 2.39) | 1.76 (1.03 – 3.01) |
| Morocco | 1.31 (0.85 – 2.01) | 1.88 (1.19 – 2.96) | 2.66 (1.65 – 4.30) |
| Suriname | 1.47 (1.06 – 2.03) | 1.82 (1.29 – 2.57) | 1.49 (1.03 – 2.15) |
| Dutch Caribbean | 0.76 (0.36 – 1.62) | 1.47 (0.68 – 3.17) | 1.39 (0.64 – 3.05) |
| Indonesia | 1.62 (1.02 – 2.58) | 1.32 (0.79 – 2.18) | 1.27 (0.76 – 2.12) |
| Other Africa | 0.83 (0.44 – 1.57) | 1.88 (0.97 – 3.62) | 2.05 (1.04 – 4.03) |
| Other Asia | 0.47 (0.25 – 0.86) | 0.82 (0.41 – 1.63) | 1.09 (0.55 – 2.16) |
| Other America’s and Oceania | 0.31 (0.10 – 0.96) | 0.94 (0.30 – 2.96) | 1.24 (0.39 – 3.89) |
| **Household Composition** |  | | |
| One person household without children | **Ref** | **-** | **Ref** |
| One parent household with children | 0.21 (0.09 – 0.47) | - | 0.62 (0.26 – 1.45) |
| Couple without children | 1.13 (0.84 – 1.52) | - | 0.91 (0.67 – 1.24) |
| Couple with children | 0.24 (0.15 – 0.37) | - | 0.85 (0.52 – 1.39) |
| More than two-person adult household without children | 0.60 (0.19 – 1.90) | - | 1.05 (0.33 – 3.32) |
| More than two-person adult household with children | 0.79 (0.34 – 1.80) | - | 1.34 (0.57 – 3.17) |
| Institutional household | 23.58 (17.71 – 31.40) | - | 17.17 (12.39 – 23.80) |
| Other household | 0.47 (0.06 – 3.34) | - | 1.43 (0.20 – 10.16) |
| **Comorbidities** |  | | |
| Neoplastic Disease | 1.77 (1.35 – 2.32) | **-** | 1.63 (1.23 – 2.17) |
| Liver Disease | 5.01 (2.88 – 8.73) | - | 2.53 (1.41 – 4.53) |
| Congestive Heart Failure | 15.99 (11.56 – 22.12) | - | 2.15 (1.50 – 3.08) |
| Cerebrovascular Disease | 10.35 (7.55 – 14.19) | - | 1.77 (1.25 – 2.51) |
| Renal Disease | 12.08 (8.20 – 17.80) | - | 1.62 (1.07 – 2.45) |
| Pulmonary Disease | 4.85 (3.56 – 6.61) | - | 1.80 (1.29 – 2.52) |
| Dementia | 20.26 (13.14 – 31.23) | - | 1.55 (0.98 – 2.46) |
| Neurologic Disease | 11.19 (7.04 – 17.80) | - | 3.83 (2.32 – 6.30) |
| Diabetes mellitus | 6.81 (5.39 – 8.61) | - | 1.63 (1.24 – 2.13) |
| HIV^d^ | - | - | - |
| Immunocompromised | 1.81 (1.15 – 2.85) | - | 1.07 (0.67 – 1.73) |
| Other Cardiovascular Disease | 6.27 (4.82 – 8.16) | - | 1.14 (0.85 – 1.54) |
| **Quartile** |  | | |
| 1 | **Ref** | **Ref** | **Ref** |
| 2 | 2.50 (1.76 – 3.55) | 2.23 (1.56 – 3.18) | 2.28 (1.59 – 3.25) |
| 3 | 0.90 (0.59 – 1.38) | 0.84 (0.54 – 1.29) | 0.87 (0.56 – 1.34) |
| 4 | 2.80 (1.99 – 3.96) | 2.58 (1.82 – 3.66) | 2.69 (1.90 – 3.82) |
| **Complicated LRTI episode in the past year** |  | | |
| No | **Ref** | **Ref** | **Ref** |
| Yes | 7.31 (2.72 – 19.60) | 1.93 (0.71 – 5.23) | 0.83 0.29 – 2.35) |

a. Univariable unadjusted incidence rate ratio **b.** Adjusted Incidence rate ratio for a multivariable model with structural determinants SES, Migration background, age, and sex, and quartile of a year, and LRTI hospitalisation in the past year **c.** Adjusted Incidence rate ratio for the full multivariable model with SES, migration background, age, sex, household composition, comorbidity, quartile of a year, and LRTI hospitalisation in the past year. **d.** HIV was not included in analysis due to insufficient cases. **e.** Those above 80 years of age were excluded from our regression analyses due to the small number of individuals over 80 years of age in the migration background groups

**Table S7 – Age group specific, crude complicated LRTI incidence, LRTI-associated mortality, and overall mortality rates per 1,000 person-years by migration background, over 18 years of age in 2014-2019.**

| **The Netherlands** | | | | | |
| --- | --- | --- | --- | --- | --- |
| **Age category** | **N** | **SES** | **complicated LRTI** | **LRTI-associated mortality** | **Overall mortality** |
| **18-49** | 98,234 (48.37%) | 59 [33-80] | 0.41 | 0.01 | 0.40 |
| **50-59** | 35,642 (17.55%) | 68 [40-87] | 1.34 | 0.13 | 2.23 |
| **60-69** | 32,467 (15.99%) | 61 [33-86] | 3.21 | 0.41 | 6.04 |
| **70-79** | 22,019 (10.84%) | 47 [26-80] | 6.29 | 1.2e | 13.00 |
| **80+** | 14,713 (7.25%) | 36 [24-68] | 15.81 | 5.4e | 55.53 |
| **Total** | 203,075 (100%) | 58 [31-82] | 2.77 | 0.62 | 6.97 |
| **Middle and Eastern Europe** | | | | | |
| **Age category** | **N** | **SES** | **complicated LRTI** | **LRTI-associated mortality** | **Overall mortality** |
| **18-49** | 17,353 (87.53%) | 28 [11-46] | 0.25 | 0.02 | 0.43 |
| **50-59** | 1,629 (8.22%) | 31 [12-53] | 0.69 | 0.00 | 1.48 |
| **60-69** | 573 (2.89%) | 32 [11-70] | 0.87 | 0.00 | 2.91 |
| **70-79** | 185 (0.93%) | 51.5 [20-87] | 6.07 | 2.6e | 8.67 |
| **80+** | 86 (0.43%) | 42 [21-86] | 11.50 | 5.7e | 70.90 |
| **Total** | 19,826 (100%) | 29 [11-47] | 0.41 | 0.07 | 0.97 |
| **‘Other Europe’** | | | | | |
| **Age category** | **N** | **SES** | **complicated LRTI** | **LRTI-associated mortality** | **Overall mortality** |
| **18-49** | 21,779 (65.28%) | 48 [17-78] | 0.34 | 0.01 | 0.32 |
| **50-59** | 4,595 (13.78%) | 65 [29-89] | 0.61 | 0.04 | 2.11 |
| **60-69** | 3,117 (9.34%) | 58 [26-87] | 2.02 | 0.32 | 6.22 |
| **70-79** | 2,516 (7.54%) | 48 [25-82] | 6.03 | 1.3e | 12.40 |
| **80+** | 1,354 (4.06%) | 41 [24-79] | 12.74 | 3.2e | 41.89 |
| **Total** | 33,361 (100%) | 50 [21-81] | 1.46 | 0.26 | 3.70 |
| **Turkey** | | | | | |
| **Age category** | **N** | **SES** | **complicated LRTI** | **LRTI-associated mortality** | **Overall mortality** |
| **18-49** | 21,733 (76.98%) | 29 [12-49] | 0.50 | 0.01 | 0.23 |
| **50-59** | 3,978 (14.09%) | 27 [12-51] | 1.64 | 0.17 | 1.14 |
| **60-69** | 1,611 (5.71%) | 17 [11-36] | 4.50 | 0.10 | 3.87 |
| **70-79** | 745 (2.64%) | 16 [11-26] | 13.32 | 0.90 | 14.68 |
| **80+** | 164 (0.58%) | 17 [11-34.5] | 18.28 | 1.9e | 23.08 |
| **Total** | 28,231 (100%) | 27 [12-48] | 1.33 | 0.07 | 1.08 |
| **Morocco** | | | | | |
| **Age category** | **N** | **SES** | **complicated LRTI** | **LRTI-associated mortality** | **Overall mortality** |
| **18-49** | 15,419 (75.42%) | 20 [9-41] | 0.51 | 0.00 | 0.32 |
| **50-59** | 2,638 (12.87%) | 16 [10-32] | 1.34 | 0.06 | 1.79 |
| **60-69** | 1,438 (5.45%) | 16 [13-25] | 4.84 | 0.35 | 3.80 |
| **70-79** | 845 (4.21%) | 16 [13-23] | 15.46 | 1.2e | 12.52 |
| **80+** | 153 (0.75%) | 15 [11-18] | 23.80 | 4.1e | 28.98 |
| **Total** | 20,493 (100%) | 18 [9-39] | 1.72 | 0.11 | 1.48 |
| **Suriname** | | | | | |
| **Age category** | **N** | **SES** | **complicated LRTI** | **LRTI-associated mortality** | **Overall mortality** |
| **18-49** | 23,264 (59.90%) | 42 [21-64] | 0.81 | 0.01 | 0.54 |
| **50-59** | 7,831 (20.16%) | 46 [21-69] | 1.69 | 0.09 | 1.67 |
| **60-69** | 4,931 (12.70%) | 39 [17-65] | 4.27 | 0.44 | 5.74 |
| **70-79** | 2,150 (5.54%) | 22 [15-46] | 10.30 | 1.1e | 11.48 |
| **80+** | 659 (1.70%) | 16 [14-27] | 16.17 | 3.2e | 38.80 |
| **Total** | 38,835 (100%) | 41 [18-64] | 2.21 | 0.20 | 2.69 |
| **Dutch Caribbean** | | | | | |
| **Age category** | **N** | **SES** | **complicated LRTI** | **LRTI-associated mortality** | **Overall mortality** |
| **18-49** | 7,407 (75.08%) | 18 [4-44] | 0.47 | 0.00 | 0.31 |
| **50-59** | 1,271 (12.88%) | 30 [12-56] | 0.65 | 0.00 | 2.21 |
| **60-69** | 766 (7.76%) | 26 [10-53] | 2.97 | 0.64 | 5.31 |
| **70-79** | 342 (3.47%) | 18 [7-45.5] | 5.91 | 0.00 | 11.33 |
| **80+** | 80 (0.81%) | 22 [6-59] | 12.11 | 2.0e | 28.26 |
| **Total** | 9,866 (100%) | 20 [6-47] | 0.97 | 0.07 | 1.55 |
| **Indonesia** | | | | | |
| **Age category** | **N** | **SES** | **complicated LRTI** | **LRTI-associated mortality** | **Overall mortality** |
| **18-49** | 6,536 (38.22% | 57 [31-79] | 0.36 | 0.00 | 0.59 |
| **50-59** | 3,765 (22.02%) | 68 [36-87] | 0.93 | 0.04 | 1.60 |
| **60-69** | 3,607 (21.09%) | 66 [36-89] | 1.86 | 0.33 | 3.68 |
| **70-79** | 1,743 (10.19%) | 59 [31-90] | 4.66 | 0.90 | 12.38 |
| **80+** | 1,449 (8.47%) | 51 [32-84] | 13.83 | 4.4e | 55.55 |
| **Total** | 17,100 (100%) | 61 [33-84] | 2.40 | 0.55 | 7.38 |
| **‘Other Africa’** | | | | | |
| **Age category** | **N** | **SES** | **complicated LRTI** | **LRTI-associated mortality** | **Overall mortality** |
| **18-49** | 9,353 (74.12%) | 17 [8-40] | 0.42 | 0.00 | 0.32 |
| **50-59** | 2,306 (18.28%) | 21 [10-43] | 1.17 | 0.00 | 1.02 |
| **60-69** | 726 (5.75%) | 19 [10-42] | 1.83 | 0.00 | 3.65 |
| **70-79** | 173 (1.37%) | 17 [13-44] | 3.70 | 0.00 | 6.47 |
| **80+** | 60 (0.48%) | 32 [14-79] | 12.11 | 0.00 | 21.20 |
| **Total** | 12,618 (100%) | 18 [9-41] | 0.74 | 0.00 | 0.82 |
| **‘Other Asia’** | | | | | |
| **Age category** | **N** | **SES** | **complicated LRTI** | **LRTI-associated mortality** | **Overall mortality** |
| **18-49** | 17,122 (76.00%) | 29 [9-58] | 0.31 | 0.00 | 0.23 |
| **50-59** | 2,904 (12.89%) | 28 [11-57] | 0.86 | 0.06 | 1.03 |
| **60-69** | 1,718 (7.63%) | 21 [11-53] | 3.10 | 0.48 | 4.56 |
| **70-79** | 570 (2.53%) | 16 [13-42] | 5.69 | 1.7e | 11.66 |
| **80+** | 214 (0.95%) | 14 [11-35] | 18.11 | 5.5e | 34.64 |
| **Total** | 22,528 (100%) | 28 [10-57] | 0.90 | 0.14 | 1.28 |
| **‘Other America and Oceania’** | | | | | |
| **Age category** | **N** | **SES** | **complicated LRTI** | **LRTI-associated mortality** | **Overall mortality** |
| **18-49** | 7,560 (75.61%) | 36 [10-71] | 0.20 | 0.02 | 0.28 |
| **50-59** | 1,389 (13.89%) | 47 [14-85] | 1.19 | 0.24 | 1.19 |
| **60-69** | 773 (7.73%) | 37 [12-81] | 1.32 | 0.22 | 3.53 |
| **70-79** | 208 (2.08%) | 37 [13-87] | 8.48 | 1.5e | 10.79 |
| **80+** | 69 (0.69%) | 47 [14-91] | 10.13 | 0.00 | 35.45 |
| **Total** | 9,999 (100%) | 38 [11-74] | 0.66 | 0.10 | 1.11 |

Population at 1 January 2017 was used as reference for the number per age category and SES. N shown as the number of individuals belonging to the specific age category at 1 January 2017, with (%) of the total amount of individuals in the migration background population. SES in median with interquartile range [IQR]. Complicated LRTI, LRTI-associated mortality and overall mortality are shown as crude incidence rates per 1,000 person years from 2014-2019. 13 missing values regarding migration background (0.003%), 7,567 regarding SES (1.84%).

**Table S8 – Age group specific, crude complicated LRTI incidence, LRTI-associated mortality, and overall mortality rates per 1,000 person-years by migration background, over 18 years of age in 2020.**

| **The Netherlands** | | | | | |
| --- | --- | --- | --- | --- | --- |
| **Age category** | **N** | **SES** | **complicated LRTI** | **LRTI-associated mortality** | **Overall mortality** |
| **18-49** | 92,158 (46.41%) | 60 [34-81] | 0.56 | 0.05 | 1.08 |
| **50-59** | 35,571 (17.91%) | 71 [42-89] | 2.23 | 0.17 | 4.12 |
| **60-69** | 31,774 (16.00%) | 64 [34-88] | 5.73 | 1.2e | 14.23 |
| **70-79** | 24,720 (12.45%) | 49 [26-81] | 10.43 | 4.2e | 31.85 |
| **80+** | 14,367 (7.23%) | 35 [22-69] | 37.99 | 28e | 150e |
| **Total** | 198,590 (100%) | 60 [32-84] | 5.64 | 2.78 | 18.15 |
| **Middle and Eastern Europe** | | | | | |
| **Age category** | **N** | **SES** | **complicated LRTI** | **LRTI-associated mortality** | **Overall mortality** |
| **18-49** | 22,132 (85.29%) | 33 [15-51] | 0.62 | 0.09 | 1.23 |
| **50-59** | 2,595 (10.00%) | 34 [14-54] | 2.16 | 0.00 | 3.97 |
| **60-69** | 833 (3.21%) | 24 [10-61] | 5.66 | 1.1e | 16.99 |
| **70-79** | 281 (1.08%) | 52 [18-85.5] | 3.44 | 0.00 | 44.71 |
| **80+** | 109 (0.42%) | 36.5 [18-70] | 17.72 | 18e | 120e |
| **Total** | 25,950 (100%) | 33 [14-52] | 1.05 | 0.19 | 3.03 |
| **‘Other Europe’** | | | | | |
| **Age category** | **N** | **SES** | **complicated LRTI** | **LRTI-associated mortality** | **Overall mortality** |
| **18-49** | 24,276 (66.44%) | 49 [19-78] | 0.37 | 0.00 | 0.41 |
| **50-59** | 5,147 (14.09%) | 69 [35-90] | 2.81 | 0.75 | 3.56 |
| **60-69** | 3,053 (8.36%) | 62 [28-90] | 4.92 | 1.6e | 10.50 |
| **70-79** | 2,489 (6.81%) | 50 [24-83] | 12.78 | 5.2e | 29.95 |
| **80+** | 1,571 (4.30%) | 40 [22-76] | 34.75 | 25e | 120e |
| **Total** | 36,536 (100%) | 52 [23-82] | 3.42 | 1.65 | 8.95 |
| **Turkey** | | | | | |
| **Age category** | **N** | **SES** | **complicated LRTI** | **LRTI-associated mortality** | **Overall mortality** |
| **18-49** | 22,630 (75.14%) | 35 [17-56] | 2.17 | 0.09 | 0.44 |
| **50-59** | 4,442 (14.75%) | 32.5 [12-56] | 11.27 | 1.1e | 4.55 |
| **60-69** | 1,965 (6.52%) | 17 [10-40.5] | 23.39 | 3.0e | 13.44 |
| **70-79** | 813 (2.70%) | 14 [10-28] | 40.79 | 7.9e | 31.73 |
| **80+** | 265 (0.88%) | 15 [10-34] | 62.36 | 38e | 110e |
| **Total** | 30,115 (100%) | 33 [14-55] | 6.65 | 1.02 | 3.92 |
| **Morocco** | | | | | |
| **Age category** | **N** | **SES** | **complicated LRTI** | **LRTI-associated mortality** | **Overall mortality** |
| **18-49** | 16,152 (73.10%) | 24 [10-45] | 2.55 | 0.06 | 0.81 |
| **50-59** | 3,029 (13.71%) | 18 [9-38] | 9.50 | 0.32 | 1.58 |
| **60-69** | 1,689 (7.65%) | 14 [10-26] | 31.10 | 4.7e | 11.74 |
| **70-79** | 959 (4.34%) | 14 [11-20] | 45.25 | 14e | 37.02 |
| **80+** | 266 (1.20%) | 13 [10-16] | 53.86 | 6.3e | 38.02 |
| **Total** | 22,095 (100%) | 20 [10-42] | 8.32 | 1.17 | 3.87 |
| **Suriname** | | | | | |
| **Age category** | **N** | **SES** | **complicated LRTI** | **LRTI-associated mortality** | **Overall mortality** |
| **18-49** | 22,025 (55.92%) | 45 [23-67] | 2.26 | 0.14 | 1.62 |
| **50-59** | 8,050 (20.44%) | 49 [23-72] | 8.70 | 0.49 | 4.78 |
| **60-69** | 5,862 (14.88%) | 40 [16-68] | 12.82 | 1.9e | 10.46 |
| **70-79** | 2,564 (6.51%) | 25 [14-51] | 24.53 | 11e | 37.16 |
| **80+** | 884 (2.24%) | 15 [12-29] | 53.34 | 33e | 140e |
| **Total** | 39,385 (100%) | 44 [19-68] | 7.86 | 1.93 | 9.06 |
| **Dutch Caribbean** | | | | | |
| **Age category** | **N** | **SES** | **complicated LRTI** | **LRTI-associated mortality** | **Overall mortality** |
| **18-49** | 8,123 (73.98%) | 21 [5-45] | 1.57 | 0.12 | 0.85 |
| **50-59** | 1,390 (12.66%) | 32 [12-61] | 4.95 | 0.00 | 3.54 |
| **60-69** | 926 (8.43%) | 24 [10-53] | 5.25 | 1.0e | 16.79 |
| **70-79** | 418 (3.81%) | 20 [7-46] | 23.01 | 9.2e | 34.52 |
| **80+** | 123 (1.12%) | 13 [5-33] | 40.10 | 24e | 140e |
| **Total** | 10,980 (100%) | 23 [7-49] | 3.57 | 0.80 | 5.44 |
| **Indonesia** | | | | | |
| **Age category** | **N** | **SES** | **complicated LRTI** | **LRTI-associated mortality** | **Overall mortality** |
| **18-49** | 5,590 (33.98%) | 57 [31-80] | 0.92 | 0.18 | 0.55 |
| **50-59** | 3,661 (22.25%) | 70 [40-90] | 4.09 | 0.00 | 4.36 |
| **60-69** | 3,533 (21.48%) | 67 [36-90] | 7.16 | 2.0e | 10.31 |
| **70-79** | 2,241 (13.62%) | 62.5 [32-90] | 11.68 | 4.8e | 27.25 |
| **80+** | 1,426 (8.67%) | 49 [29-82] | 35.55 | 31e | 150e |
| **Total** | 16,454 (100%) | 62 [34-86] | 7.54 | 3.86 | 20.17 |
| **‘Other Africa’** | | | | | |
| **Age category** | **N** | **SES** | **complicated LRTI** | **LRTI-associated mortality** | **Overall mortality** |
| **18-49** | 10,460 (72.78%) | 22 [8-43] | 1.62 | 0.00 | 1.05 |
| **50-59** | 2,503 (17.41%) | 24 [10-45] | 7.05 | 1.6e | 3.92 |
| **60-69** | 1,093 (7.60%) | 19 [10-43] | 9.32 | 1.7e | 5.08 |
| **70-79** | 247 (1.72%) | 15 [12-40] | 45.61 | 15e | 38.01 |
| **80+** | 70 (0.49%) | 20 [13-66] | 43.24 | 29e | 130e |
| **Total** | 14,373 (100%) | 21 [9-44] | 4.19 | 0.82 | 3.16 |
| **‘Other Asia’** | | | | | |
| **Age category** | **N** | **SES** | **complicated LRTI** | **LRTI-associated mortality** | **Overall mortality** |
| **18-49** | 21,390 (75.68%) | 33 [11-63] | 0.98 | 0.05 | 0.61 |
| **50-59** | 3,595 (12.72% | 31 [11-62] | 5.37 | 0.54 | 4.30 |
| **60-69** | 2,199 (7.78%) | 21 [10-56] | 6.30 | 0.45 | 8.55 |
| **70-79** | 787 (2.78%) | 15 [11-42] | 15.29 | 8.2e | 22.35 |
| **80+** | 291 (1.03%) | 12 [9-29] | 55.41 | 45e | 160e |
| **Total** | 28,262 (100%) | 31 [10-61] | 2.95 | 0.84 | 4.00 |
| **‘Other America and Oceania’** | | | | | |
| **Age category** | **N** | **SES** | **complicated LRTI** | **LRTI-associated mortality** | **Overall mortality** |
| **18-49** | 8,702 (75.02%) | 39 [12-71] | 0.46 | 0.00 | 0.57 |
| **50-59** | 1,586 (13.67%) | 46 [16-83.5] | 2.99 | 0.00 | 1.79 |
| **60-69** | 917 (7.91%) | 36.5 [11-79] | 6.39 | 1.1e | 3.19 |
| **70-79** | 314 (2.71%) | 32.5 [12-81] | 18.04 | 6.0e | 21.05 |
| **80+** | 80 (0.69%) | 34 [11-85] | 11.66 | 12e | 81.65 |
| **Total** | 11,599 (100%) | 40 [12-74] | 1.86 | 0.34 | 2.11 |

Population at 1 January 2020 was used as reference for the number per age category and SES. N shown as the number of individuals belonging to the specific age category at 1 January 2020, with (%) of the total amount of individuals in the migration background population. SES in median with interquartile range [IQR]. Complicated LRTI, LRTI-associated mortality and overall mortality are shown as crude incidence rates per 1,000 person years in 2020. 8,071 values missing regarding SES (1.86%).

**Table S9 – SES-quintile specific, crude complicated LRTI incidence, LRTI-associated mortality, and overall mortality rates per 1,000 person-years by migration background, over 18 years of age in 2014-2019.**

| **The Netherlands** | | | | | |
| --- | --- | --- | --- | --- | --- |
| **SES quintile** | **N** | **Age** | **complicated LRTI** | **LRTI-associated mortality** | **Overall mortality** |
| **1** | 53,422 (26.55%) | 53 [41-64] | 1.40 | 0.22 | 3.81 |
| **2** | 42,789 (21.27%) | 48 [35-60] | 1.45 | 0.27 | 3.42 |
| **3** | 37,554 (18.66%) | 48 [34-66] | 2.16 | 0.42 | 5.08 |
| **4** | 35,897 (17.84%) | 58 [37-73] | 4.52 | 1.0e | 10.10 |
| **5** | 31,552 (15.68%) | 48 [28-64] | 4.46 | 1.0e | 8.74 |
| **Middle and Eastern Europe** | | | | | |
| **SES quintile** | **N** | **Age** | **complicated LRTI** | **LRTI-associated mortality** | **Overall mortality** |
| **1** | 1,051 (5.50%) | 39 [29-49] | 0.49 | 0.16 | 3.28 |
| **2** | 1,682 (8.81%) | 37 [30-45] | 0.10 | 0.00 | 0.67 |
| **3** | 3,495 (18.30%) | 35 [29-42] | 0.38 | 0.05 | 0.72 |
| **4** | 5,372 (28.13%) | 35 [29-42] | 0.35 | 0.06 | 0.48 |
| **5** | 7,497 (39.26%) | 33 [26-42] | 0.47 | 0.07 | 0.90 |
| **Other Europe** | | | | | |
| **SES quintile** | **N** | **Age** | **complicated LRTI** | **LRTI-associated mortality** | **Overall mortality** |
| **1** | 8,165 (25.92%) | 48 [39-59] | 1.10 | 0.14 | 3.10 |
| **2** | 5,156 (16.37%) | 43 [34-55] | 0.99 | 0.19 | 2.43 |
| **3** | 4,987 (15.83%) | 43 [33-56] | 1.10 | 0.17 | 2.83 |
| **4** | 5,354 (16.99%) | 43 [32-65] | 2.15 | 0.44 | 4.43 |
| **5** | 7,842 (24.89%) | 36 [25-53] | 1.85 | 0.26 | 3.79 |
| **Turkey** | | | | | |
| **SES quintile** | **N** | **Age** | **complicated LRTI** | **LRTI-associated mortality** | **Overall mortality** |
| **1** | 1,044 (3.72%) | 40 [31-48] | 0.00 | 0.00 | 0.46 |
| **2** | 3,045 (10.85%) | 37 [28-48] | 0.83 | 0.06 | 0.89 |
| **3** | 4,885 (17.41%) | 36 [27-47] | 0.99 | 0.00 | 0.82 |
| **4** | 7,476 (26.64%) | 37 [28-47] | 0.99 | 0.09 | 0.83 |
| **5** | 11,611 (41.38%) | 39 [29-52] | 1.95 | 0.10 | 1.40 |
| **Morocco** | | | | | |
| **SES quintile** | **N** | **Age** | **complicated LRTI** | **LRTI-associated mortality** | **Overall mortality** |
| **1** | 498 (2.44%) | 38 [31-46] | 0.98 | 0.00 | 0.00 |
| **2** | 1,428 (7.00%) | 35 [28-45] | 0.35 | 0.12 | 0.58 |
| **3** | 2,764 (13.55%) | 35 [27-45] | 1.17 | 0.06 | 1.04 |
| **4** | 4,654 (22.81%) | 36 [27-49] | 1.22 | 0.00 | 1.22 |
| **5** | 11,060 (54.21%) | 40 [30-53] | 2.30 | 0.18 | 1.84 |
| **Suriname** | | | | | |
| **SES quintile** | **N** | **Age** | **complicated LRTI** | **LRTI-associated mortality** | **Overall mortality** |
| **1** | 3,843 (9.97%) | 48 [34-58] | 1.22 | 0.17 | 1.35 |
| **2** | 7,346 (19.06%) | 45 [32-54] | 1.02 | 0.07 | 1.16 |
| **3** | 8,407 (21.81%) | 44 [32-54] | 1.55 | 0.06 | 1.06 |
| **4** | 8,370 (21.71%) | 44 [31-56] | 1.97 | 0.20 | 2.53 |
| **5** | 10,582 (27.45%) | 48 [32-62] | 3.89 | 0.36 | 4.77 |
| **Dutch Caribbean** | | | | | |
| **SES quintile** | **N** | **Age** | **complicated LRTI** | **LRTI-associated mortality** | **Overall mortality** |
| **1** | 591 (6.05%) | 45 [35-55] | 0.93 | 0.00 | 1.54 |
| **2** | 927 (9.48%) | 40 [32-52] | 0.50 | 0.17 | 1.18 |
| **3** | 1,403 (14.35%) | 39 [30-52] | 0.24 | 0.00 | 0.84 |
| **4** | 1,925 (19.69%) | 36 [27-51] | 0.77 | 0.00 | 1.03 |
| **5** | 4,930 (50.43%) | 30 [24-47] | 1.27 | 0.07 | 1.41 |
| **Indonesia** | | | | | |
| **SES quintile** | **N** | **Age** | **complicated LRTI** | **LRTI-associated mortality** | **Overall mortality** |
| **1** | 5,050 (30.07%) | 58 [48-68] | 1.80 | 0.47 | 5.84 |
| **2** | 3,463 (20.62%) | 53 [42-64] | 1.50 | 0.14 | 4.49 |
| **3** | 2,952 (17.58%) | 54 [41-67] | 2.87 | 0.72 | 7.62 |
| **4** | 2,818 (16.78%) | 58 [43-72] | 3.51 | 0.85 | 9.70 |
| **5** | 2,511 (14.95%) | 52 [37-63] | 2.37 | 0.26 | 4.41 |
| **‘Other Africa’** | | | | | |
| **SES quintile** | **N** | **Age** | **complicated LRTI** | **LRTI-associated mortality** | **Overall mortality** |
| **1** | 786 (6.44%) | 43 [32-53] | 0.45 | 0.00 | 1.56 |
| **2** | 871 (7.13%) | 39 [30-51] | 0.54 | 0.00 | 0.54 |
| **3** | 1,434 (11.75%) | 39 [30-52] | 0.57 | 0.00 | 0.80 |
| **4** | 2,569 (21.04%) | 40 [29-51] | 0.77 | 0.00 | 0.77 |
| **5** | 6,548 (53.64%) | 37 [27-50] | 0.83 | 0.00 | 0.70 |
| **‘Other Asia’** | | | | | |
| **SES quintile** | **N** | **Age** | **complicated LRTI** | **LRTI-associated mortality** | **Overall mortality** |
| **1** | 2,487 (11.63%) | 39 [33-50] | 0.41 | 0.28 | 1.03 |
| **2** | 2,429 (11.36%) | 37 [31-47] | 0.55 | 0.07 | 0.62 |
| **3** | 3,043 (14.23%) | 37 [30-47] | 0.49 | 0.11 | 1.08 |
| **4** | 4,194 (19.61%) | 37 [29-49] | 0.80 | 0.04 | 0.68 |
| **5** | 9,233 (43.17%) | 37 [28-52] | 1.29 | 0.13 | 1.67 |
| **‘Other America and Oceania’** | | | | | |
| **SES quintile** | **N** | **Age** | **complicated LRTI** | **LRTI-associated mortality** | **Overall mortality** |
| **1** | 1,895 (20.41%) | 44 [35-54] | 0.81 | 0.00 | 1.17 |
| **2** | 1,261 (13.58%) | 38 [31-48] | 0.66 | 0.13 | 0.53 |
| **3** | 1,291 (13.91%) | 39 [30-49] | 0.13 | 0.13 | 0.64 |
| **4** | 1,505 (16.21%) | 38 [28-49] | 0.56 | 0.00 | 1.00 |
| **5** | 3,331 (35.88%) | 35 [26-50] | 0.95 | 0.15 | 1.40 |

Population at 1 January 2017 was used as reference for the number per SES quintile. N shown as the number of individuals belonging to the specific SES quintile at 1 January 2017, with (%) of the total amount of individuals in the migration background population. Age shown as median with interquartile range [IQR]. SES in quintiles where 5 is the lowest quintile. Complicated LRTI, LRTI-associated mortality and overall mortality are shown as crude incidence rates per 1,000 person years from 2014-2019. 7,657 missing observations regarding SES (1.84%), 13 missing observations regarding migration background (0.003%)

**Table S10 – SES-quintile specific, crude complicated LRTI incidence, LRTI-associated mortality, and overall mortality rates per 1,000 person-years by migration background, over 18 years of age in 2020.**

| **The Netherlands** | | | | | |
| --- | --- | --- | --- | --- | --- |
| **SES quintile** | **N** | **Age** | **complicated LRTI** | **LRTI-associated mortality** | **Overall mortality** |
| **1** | 56,990 (28.86%) | 53 [41-65] | 2.96 | 1.12 | 8.73 |
| **2** | 41,174 (20.85%) | 49 [35-62] | 3.76 | 1.32 | 10.64 |
| **3** | 35,164 (17.81%) | 49 [34-65] | 4.62 | 2.09 | 14.91 |
| **4** | 34,064 (17.25%) | 58 [36-73] | 9.01 | 5.23 | 31.93 |
| **5** | 30,054 (15.22%) | 52 [31-68] | 9.06 | 4.68 | 28.84 |
| **Middle and Eastern Europe** | | | | | |
| **SES quintile** | **N** | **Age** | **complicated LRTI** | **LRTI-associated mortality** | **Overall mortality** |
| **1** | 1,440 (5.77%) | 43 [35-52] | 1.40 | 0.00 | 4.20 |
| **2** | 2,836 (11.36%) | 37 [31-45] | 1.08 | 0.00 | 2.88 |
| **3** | 5,385 (21.56%) | 36 [30-45] | 0.38 | 0.00 | 1.72 |
| **4** | 6,898 (27.62%) | 36 [30 -43] | 1.48 | 0.45 | 2.67 |
| **5** | 8,415 (33.70%) | 35 [27-45] | 1.24 | 0.12 | 3.36 |
| **Other Europe** | | | | | |
| **SES quintile** | **N** | **Age** | **complicated LRTI** | **LRTI-associated mortality** | **Overall mortality** |
| **1** | 9,100 (26.71%) | 48 [39-58] | 2.56 | 1.22 | 6.45 |
| **2** | 5,749 (16.87%) | 43 [33-55] | 3.37 | 1.42 | 6.22 |
| **3** | 5,452 (16.00%) | 42 [32-55] | 3.18 | 1.50 | 8.99 |
| **4** | 5,776 (16.95%) | 42 [30-59] | 2.82 | 1.06 | 11.62 |
| **5** | 7,998 (23.47%) | 34 [25-53] | 5.62 | 2.87 | 13.72 |
| **Turkey** | | | | | |
| **SES quintile** | **N** | **Age** | **complicated LRTI** | **LRTI-associated mortality** | **Overall mortality** |
| **1** | 1,899 (6.34%) | 40 [30-50] | 7.88 | 1.58 | 2.63 |
| **2** | 4,080 (13.63%) | 37 [28-48] | 4.42 | 0.00 | 1.97 |
| **3** | 6,114 (20.53% | 37 [28-47] | 4.55 | 0.33 | 1.63 |
| **4** | 7,552 (25.23%) | 38 [28-48] | 5.14 | 0.53 | 2.63 |
| **5** | 10,257 (34.27%) | 42 [32-56] | 9.82 | 2.04 | 7.19 |
| **Morocco** | | | | | |
| **SES quintile** | **N** | **Age** | **complicated LRTI** | **LRTI-associated mortality** | **Overall mortality** |
| **1** | 760 (3.46%) | 38 [29-47] | 3.95 | 0.00 | 2.63 |
| **2** | 1,964 (8.93%) | 35 [28-46] | 2.55 | 1.02 | 2.04 |
| **3** | 3,172 (14.43%) | 36 [27-46] | 6.63 | 0.95 | 2.52 |
| **4** | 5,087 (23.13%) | 36 [26-49] | 6.66 | 0.00 | 1.96 |
| **5** | 11,006 (50.05%) | 42 [ 31-56] | 11e | 1.90 | 5.33 |
| **Suriname** | | | | | |
| **SES quintile** | **N** | **Age** | **complicated LRTI** | **LRTI-associated mortality** | **Overall mortality** |
| **1** | 4,905 (12.54%) | 49 [33-59] | 5.75 | 0.21 | 2.87 |
| **2** | 7,725 (19.75%) | 46 [32-57] | 6.09 | 0.65 | 3.37 |
| **3** | 8,372 (21.40%) | 45 [33-56] | 7.52 | 1.55 | 4.78 |
| **4** | 7,978 (20.40%) | 46 [33-59] | 7.69 | 1.39 | 8.58 |
| **5** | 10,134 (25.91%) | 51 [35-65] | 11e | 4.28 | 18.90 |
| **Dutch Caribbean** | | | | | |
| **SES quintile** | **N** | **Age** | **complicated LRTI** | **LRTI-associated mortality** | **Overall mortality** |
| **1** | 630 (5.80%) | 47 [36-56] | 3.11 | 1.56 | 7.78 |
| **2** | 1,215 (11.18%) | 40 [31-52] | 4.11 | 0.00 | 1.64 |
| **3** | 1,585 (14.59%) | 39 [30-52] | 5.69 | 0.63 | 2.53 |
| **4** | 2,220 (20.44%) | 35 [28-50] | 3.18 | 0.45 | 4.09 |
| **5** | 5,213 (47.99%) | 29 [24-49] | 3.07 | 1.15 | 6.90 |
| **Indonesia** | | | | | |
| **SES quintile** | **N** | **Age** | **complicated LRTI** | **LRTI-associated mortality** | **Overall mortality** |
| **1** | 5,181 (31.91%) | 59 [50-69] | 5.63 | 1.94 | 13.00 |
| **2** | 3,228 (19.88%) | 55 [43-67] | 5.32 | 4.07 | 13.15 |
| **3** | 2,782 (17.14%) | 55 [42-68] | 5.83 | 2.19 | 21.14 |
| **4** | 2,665 (16.42%) | 60 [45-73] | 12e | 6.46 | 35.37 |
| **5** | 2,379 (14.65%) | 56 [39-67] | 9.79 | 4.25 | 20.85 |
| **‘Other Africa’** | | | | | |
| **SES quintile** | **N** | **Age** | **complicated LRTI** | **LRTI-associated mortality** | **Overall mortality** |
| **1** | 922 (6.63%) | 44 [33-53] | 4.41 | 0.00 | 2.21 |
| **2** | 1,210 (8.70%) | 38 [29-50] | 3.34 | 0.84 | 2.51 |
| **3** | 1,760 (12.66%) | 39 [29-52] | 3.99 | 0.57 | 5.13 |
| **4** | 3,246 (23.34%) | 38 [27-51] | 2.76 | 0.00 | 2.15 |
| **5** | 6,769 (48.67%) | 38 [28-52] | 5.47 | 1.48 | 3.69 |
| **‘Other Asia’** | | | | | |
| **SES quintile** | **N** | **Age** | **complicated LRTI** | **LRTI-associated mortality** | **Overall mortality** |
| **1** | 3,418 (12.73%) | 41 [34-51] | 0.89 | 0.00 | 1.78 |
| **2** | 3,409 (12.70%) | 37 [31-47] | 1.49 | 0.59 | 2.67 |
| **3** | 3,939 (14.67%) | 38 [30-48] | 2.81 | 0.51 | 2.30 |
| **4** | 5,338 (19.88%) | 37 [29-48] | 2.45 | 0.00 | 1.32 |
| **5** | 10,742 (40.01%) | 38 [29-53] | 4.51 | 1.69 | 7.05 |
| **‘Other America and Oceania’** | | | | | |
| **SES quintile** | **N** | **Age** | **complicated LRTI** | **LRTI-associated mortality** | **Overall mortality** |
| **1** | 2,200 (20.21%) | 44 [35-54] | 0.00 | 0.00 | 1.38 |
| **2** | 1,564 (14.37%) | 38 [31-49] | 1.93 | 0.00 | 1.29 |
| **3** | 1,609 (14.78%) | 38 [30-48] | 1.89 | 0.00 | 0.00 |
| **4** | 1,832 (16.83%) | 37 [29-49] | 0.55 | 0.00 | 1.10 |
| **5** | 3,679 (33.80%) | 35 [26-51] | 4.12 | 1.10 | 4.67 |

Population at 1 January 2020 was used as reference for the number per SES quintile. N shown as the number of individuals belonging to the specific SES quintile at 1 January 2020, with (%) of the total amount of individuals in the migration background population. Age shown as median with interquartile range [IQR]. SES in quintiles where 5 is the lowest quintile. Complicated LRTI, LRTI-associated mortality and overall mortality are shown as crude incidence rates per 1,000 person years in 2020. 8,071 observations missing regarding SES (1.86%).

**Table S11 – SES-quintile specific, crude complicated LRTI incidence rates per 1,000 person-years, by flu season, over 18 years of age from 2014 through 2019.**

|  | **Flu season** | | | | | | |
| --- | --- | --- | --- | --- | --- | --- | --- |
| **SES quintile** | **13/14** | **14/15** | **15/16** | **16/17** | **17/18** | **18/19** | **19/20** |
| 1 | 1.55 | 1.58 | 0.96 | 1.44 | 1.20 | 1.37 | 0.93 |
| 2 | 1.32 | 1.12 | 1.04 | 1.21 | 1.42 | 1.33 | 1.10 |
| 3 | 1.64 | 1.55 | 1.51 | 1.60 | 1.85 | 1.80 | 1.71 |
| 4 | 2.26 | 2.95 | 2.19 | 2.70 | 3.19 | 3.04 | 2.98 |
| 5 | 2.32 | 2.46 | 2.01 | 2.42 | 3.08 | 3.19 | 2.87 |

Flu season was defined based on yearly quartiles where a flu season ranged from 1 July until 30 June the following year. Flu season 13/14 encompassed 1 January 2014 until 30 June 2014, flu season 19/20 encompassed 1 July 2019 until 31 December 2019. SES categorised in quintiles where 5 is the lowest. Table shows crude complicated LRTI incidence rates per 1,000 person-years per flu-season from 1 January 2014 until 31 December 2019. 7,657 missing observations regarding SES (1.84%)

**Table S12 – Fully adjusted multivariable Poisson regression sensitivity analysis regarding LRTI-associated mortality, which includes deaths occurring abroad in the composite outcome measure, between the ages of 18 and 80, in 2014-2019.**

|  | **Jan 2014 – Dec 2019** | |
| --- | --- | --- |
| **Independent Variables** | **aIRR^1^ (95% CI)** | **P-value** |
| **Sex** |  | |
| Male | **Ref** | **Ref** |
| Female | 0.61 (0.50 – 0.76) | 0.000 |
| **Age** |  | |
| 18-49 | 0.02 (0.01 – 0.04) | 0.000 |
| 50 – 59 | 0.14 (0.10 – 0.20) | 0.000 |
| 60 – 69 | 0.41 (0.32 – 0.52) | 0.000 |
| 70 – 80 | **Ref** | **Ref** |
| **SES (quintiles where 5 is the lowest)** |  | |
| 1 | **Ref** | **Ref** |
| 2 | 1.21 (0.77 – 1.89) | 0.405 |
| 3 | 2.01 (1.35 – 2.99) | 0.001 |
| 4 | 2.09 (1.43 – 3.06) | 0.000 |
| 5 | 3.07 (2.10 – 4.49) | 0.000 |
| **Migration Background** |  | |
| The Netherlands | **Ref** | **Ref** |
| Middle & Eastern Europe | 0.68 (0.25 – 1.86) | 0.453 |
| Other Europe | 0.93 (0.63 – 1.38) | 0.731 |
| Turkey | 0.80 (0.48 – 1.34) | 0.400 |
| Morocco | 1.01 (0.61 – 1.66) | 0.966 |
| Suriname | 0.90 (0.63 – 1.28) | 0.563 |
| Dutch Caribbean | 0.24 (0.06 – 0.97) | 0.045 |
| Indonesia | 0.71 (0.43 – 1.17) | 0.181 |
| Other Africa^b^ | - | - |
| Other Asia | 0.95 (0.54 – 1.67) | 0.850 |
| Other America’s and Oceania | 1.24 (0.55 – 2.82) | 0.603 |
| **Household Composition** |  | |
| One person household without children | **Ref** | **Ref** |
| One parent household with children | 0.64 (0.33 – 1.22) | 0.176 |
| Couple without children | 0.84 (0.67 – 1.07) | 0.157 |
| Couple with children | 0.71 (0.47 – 1.07) | 0.105 |
| More than two-person adult household without children | 1.51 (0.67 – 3.39) | 0.322 |
| More than two-person adult household with children | 1.60 (0.83 – 3.08) | 0.165 |
| Institutional household | 7.59 (5.48 – 10.52) | 0.000 |
| Other household | 0.78 (0.11 – 5.58) | 0.803 |
| **Comorbidities** |  | |
| Neoplastic Disease | 1.59 (1.27 – 1.99) | 0.000 |
| Liver Disease | 2.08 (1.22 – 3.53) | 0.007 |
| Congestive Heart Failure | 2.73 (2.03 – 3.66) | 0.000 |
| Cerebrovascular Disease | 0.84 (0.57 – 1.23) | 0.360 |
| Renal Disease | 1.37 (0.94 – 2.00) | 0.106 |
| Pulmonary Disease | 4.19 (3.35 – 5.23) | 0.000 |
| Dementia | 0.99 (0.59 – 1.68) | 0.984 |
| Neurologic Disease | 2.61 (1.61 – 4.24) | 0.000 |
| Diabetes mellitus | 1.08 (0.85 – 1.38) | 0.524 |
| HIV | 2.08 (0.51 – 8.40) | 0.306 |
| Immunocompromised | 0.99 (0.66 – 1.48) | 0.947 |
| Other Cardiovascular Disease | 0.83 (0.63 – 1.08) | 0.161 |
| **Flu season** |  | |
| 13/14 | **Ref** | **Ref** |
| 14/15 | 0.55 (0.39 – 0.78) | 0.001 |
| 15/16 | 0.36 (0.24 – 0.54) | 0.000 |
| 16/17 | 0.36 (0.24 – 0.52) | 0.000 |
| 17/18 | 0.46 (0.32 – 0.66) | 0.000 |
| 18/19 | 0.78 (0.56 – 1.09) | 0.152 |
| 19/20 | 1.14 (0.73 – 1.78) | 0.567 |
| **Quartile** |  | |
| 1 | **Ref** | **Ref** |
| 2 | 0.49 (0.38 – 0.63) | 0.000 |
| 3 | 0.39 (0.29 – 0.52) | 0.000 |
| 4 | 0.34 (0.25 – 0.46) | 0.000 |
| **Complicated LRTI episode in the past year** |  | |
| No | **Ref** | **Ref** |
| Yes | 3.78 (2.24 – 6.38) | 0.000 |

Flu season was defined based on yearly quartiles where a flu season ranged from 1 July until 30 June the following year. Flu season 13/14 encompassed 1 January 2014 until 30 June 2014, flu season 19/20 encompassed 1 July 2019 until 31 December 2019. **a.** Adjusted Incidence rate ratio for the full multivariable model with SES and Migration background adjusted for age, sex, household composition, comorbidity, quartile of a year, and presence of a complicated LRTI episode in the past year. Sensitivity analysis where we included a registered unknown mortality cause (ICD-10: R99) in the LRTI-associated mortality composite outcome. **b.** ‘Other Africa’ was not included in analysis due to insufficient cases.

**M-References**

1. The Hague Municipality. The Hague in numbers. <https://www.denhaag.nl/en/introducing-the-hague/the-hague-in-numbers/>. Date last updated: May 3 2024. Date last accessed: March 6 2024.
2. Statistics Netherlands. Microdata 2019. <https://www.cbs.nl/en-gb/our-services/customised-services-microdata/microdata-conducting-your-own-research/overview-of-all-datasets>. Date last accessed: November 26 2024.
3. Fine MJ, Auble TE, Yealy DM, et al. A prediction rule to identify low-risk patients with community-acquired pneumonia. N Engl J Med. 1997;336(4):243-50.
4. World Health Organization. Anatomical Therapeutic Chemical (ATC) Classification. <https://www.who.int/tools/atc-ddd-toolkit/atc-classification>. Date last accessed: December 4 2024
